# Sustained freedom from disease activity in secondary progressive multiple sclerosis by targeting invariant NKT cells: a phase 2 trial of OCH

**DOI:** 10.64898/2026.02.04.26345323

**Authors:** Ben JE Raveney, Tomoko Okamoto, Atsuko Kimura, Youwei Lin, Manabu Araki, Yukio Kimura, Noriko Sato, Yuko Shimizu, Yoichiro Nishida, Takanori Yokota, Norihide Maikusa, Masanori Taketsuna, Yu Okada, Takami Ishizuka, Harumasa Nakamura, Sachiko Miyake, Yuji Takahashi, Wakiro Sato, Takashi Yamamura

**Affiliations:** Department of Immunology, National Institute of Neuroscience, National Center of Neurology and Psychiatry (NCNP), Kodaira, Tokyo, Japan; Multiple Sclerosis Center, National Center Hospital, National Center of Neurology and Psychiatry (NCNP), Kodaira, Tokyo, Japan; Department of Neurology, National Center Hospital, National Center of Neurology and Psychiatry (NCNP), Kodaira, Tokyo, Japan; Department of Radiology, National Center Hospital, National Center of Neurology and Psychiatry (NCNP), Kodaira, Tokyo, Japan; Department of Neurology, Tokyo Women’s Medical University School of Medicine, Tokyo, Japan; Department of Neurology and Neurological Science, Graduate School of Medical and Dental Sciences, Institute of Science Tokyo, Tokyo, Japan; Center for Evolutionary Cognitive Sciences, Graduate School of Art and Sciences, The University of Tokyo, 3-8-1 Meguro, Tokyo, Japan; Translational Research Center for Medical Innovation, Foundation for Biomedical Research and Innovation at Kobe, Hyogo, Japan; Department of Clinical Research Support, Clinical Research & Education Promotion Division, National Center Hospital, NCNP; Department of Immunology, Juntendo University Graduate School of Medicine, Tokyo, Japan.

**Keywords:** Multiple sclerosis, progressive MS, GM-CSF, clinical trial, NKT cells, glycolipid, EAE, gut-brain axis, gut microbiome

## Abstract

Multiple sclerosis (MS) therapies primarily rely on lymphocyte depletion or trafficking blockade, carrying risks of systemic immunosuppression; however, such treatments have limited efficacy in secondary progressive multiple sclerosis (SPMS). Thus, drugs that target stage-specific inflammation without broad immunosuppression are an unmet clinical need.

In this double-blind, placebo-controlled phase II trial, 30 patients with relapsing MS received weekly oral OCH or placebo for 24 weeks. In the pre-specified SPMS subgroup (n=12), OCH achieved complete relapse prevention (p=0.0003), prolonged relapse-free survival (p=0.0079), no new lesions (0/6), with no evidence of disease activity (NEDA-3) in 5/6 patients. In comparison, for the placebo-treated group, 5/6 patients suffered relapses, 2/6 patients developed new lesions, and no placebo-treated SPMS achieved NEDA-3.

Invariant natural killer T (iNKT) cells, a regulatory lymphocyte population that is numerically and functionally impaired in MS, are a potential target for MS therapy. Glycolipid OCH is a selective iNKT cell stimulator, skewing the cytokine environment towards Th2. OCH treatment resulted in increased IL-4-producing Th cells in patient peripheral blood while decreasing pathogenic GM-CSF-producing Th cells. Parallel studies in mouse models of MS (EAE) corroborated this mechanism and further revealed that OCH activated gut iNKT cells. Disease amelioration by OCH depended on IL-4 and its efficacy was further enhanced by depletion of B cells. These data revealed the gut-brain axis mediation of progressive-stage pathology distinct from relapsing-remitting MS.

Findings from this bidirectional translational study uncover mechanistic differences between SPMS and other types of MS and highlight divergent roles for B cells and Th cells.

Furthermore, OCH exerts its therapeutic benefit via targeting mechanisms that are distinct from currently available drugs; exploiting iNKT cell regulatory potential to reprogram pathogenic T helper responses without lymphocyte depletion. The unique yet effective nature of OCH treatment positions it as an attractive future oral therapy for SPMS.

**One Sentence Summary:** The iNKT cell activating ligand OCH suppresses disease activity selectively in secondary progressive MS in a phase II clinical trial, revealing stage-specific IL-4-mediated immune cell interactions in MS pathology.

## Introduction

Multiple sclerosis (MS) is a chronic inflammatory demyelinating disease of the central nervous system (CNS) affecting over 2.8 million people worldwide, with prevalence rising sharply in recent decades (*1*). Damage to the CNS in MS patients leads to a variety of peripheral disabling symptoms that can differ in severity over time and between patients. The most common initial presentation, relapsing-remitting MS (RRMS), is characterized by episodic clinical exacerbations followed by periods of remission. Over time, a subset of patients with RRMS shift to a chronic form, secondary progressive MS (SPMS) (*2*). Some SPMS patients continue to suffer from relapses and as well as such temporary clinical exacerbations, SPMS patients suffer from chronic progressive disabling symptoms, which are now divided into relapse-associated worsening (RAW) and progression independent of relapse activity (PIRA) (*3*). For many patients, modern disease modifying therapies can be effective in disease management; and many of such treatments specifically target immune system components providing the strongest evidence of autoimmune mechanisms as causative in MS. Despite the availability of immune-targeting therapies, substantial unmet medical needs remain, particularly in SPMS, in which current treatments show limited efficacy.

The pathogenesis of MS involves dysregulated interactions among immune cell populations such as autoreactive T and B cells, leading to the formation of demyelinating neuroinflammatory lesions (*4, 5*). Both genetic susceptibility and environmental factors contribute to disease onset (*6*), yet the mechanisms by which environmental changes drive immune dysregulation remain incompletely understood. It has been suggested that MS pathology in individuals may result from a range of different pathogenic immune cells (*7*). Therefore, it is likely that such range of possible mechanisms driving pathogenesis could explain the gamut of disease course patterns and why different drugs have different efficacy for different patients (*8–11*), emphasizing an urgent need for novel therapeutic strategies targeting disease progression.

Natural killer T (NKT) cells, innate cell-like T lymphocytes that share features with innate natural killer (NK) cells, constitute a distinct immunoregulatory lymphocyte subset that bridges innate and adaptive immunity (*12*). Unlike NK cells, NKT cells are activated via T cell receptors (TcR) and unlike T cells, NKT cells have functional expression of NK cell markers and a restricted TcR repertoire (*12*). In particular, iNKT cells, a major NKT cell population with an invariant TcR (Vα24-Jα18/Vβ11), are uniquely activated by glycolipids, rather than peptides, presented in the context of CD1d molecules, rather than MHC I/II (*13, 14*). As the major source of such glycolipid antigens is suggested to be gut-resident bacteria, the interest in the involvement of iNKT cells in host immune system/gut microbiome interactions is increasing.

The synthetic glycolipid OCH (2S, 3S, 4R -1-*O*-(α-D-Galactosyl) -2-tetracosanoylamino-1, 3, 4-nonaetriol), a structural analogue of α-galactosylceramide (α-GalCer, the prototype iNKT cell ligand), selectively activates iNKT cells to proliferate and preferentially induce IL-4 while limiting pro-inflammatory cytokine production (*15, 16*). Our early studies demonstrated that oral OCH ameliorates preclinical models of autoimmune disease, including experimental autoimmune encephalomyelitis (EAE), a mouse model of MS (*16–18*). These studies were carried out at a time when Th1 and Th2 cells were the primary recognized effector T-cell subsets, prior to our current understanding of the importance of pathogenic Th17 cells and GM-CSF-producing Th cells in EAE/MS, precluding detailed mechanistic studies of OCH effects on pathogenesis.

We have previously reported the findings of a first-in-human phase I trial of OCH in healthy volunteers and RRMS patients, showing oral OCH was safe and well tolerated (*19*). Here, we now report a double-blind, randomized, placebo-controlled phase II clinical trial evaluating oral OCH (OCH-NCNP1) in patients with relapsing MS (rMS), including both RRMS and SPMS. We demonstrate that oral OCH was associated with decreased clinical relapse activity and a reduction in the proportion of pathogenic Th cell populations, including GM-CSF-producing Th cells. Efficacy of OCH was most notable in the SPMS subgroup despite the modest sample size. Complementary EAE studies in mice suggested and supported a mechanistic link between OCH administration and attenuation of GM-CSF-producing Th cells: activation of gut iNKT cells led to IL-4-mediated modulation of Th cell activation and decreasing pathogenic populations. Interestingly, the efficacy of OCH was enhanced following B cell depletion, which may explain why OCH is most effective in SPMS patients over RRMS, as RRMS is associated with activated B cells. Taken together, these findings provide initial clinical evidence that activation of iNKT cells may modulate disease-relevant immune pathways in SPMS, a subtype with historically limited therapeutic options, encouraging further investigation of OCH in larger, clinically more vigorous studies.

## Results

### Enrolled patients and safety profile

This exploratory, investigator-initiated phase II trial (UMIN000038903) enrolled 30 MS patients, diagnosed according to the revised McDonald criteria(*20*), all with relapsing MS. The cohort included 12 SPMS patients, all exhibiting PIRA (*21*) and clinical relapses, and 18 RRMS patients experiencing relapses without PIRA. PIRA was defined as a confirmed ≥3-month increase in the EDSS score in the absence of relapse (*22*). Inclusion as relapsing MS required either 1 clinical exacerbation in the 12 months prior to recruitment or 2 within 24 months; further criteria are summarised in **Supplementary Table 1**. Patients were randomised in Placebo or OCH groups, with demographics provided in **Supplementary Table 2**. Placebo or OCH oral granules were administered weekly for 24 weeks, with blinding for both patients and attending physicians.

Twenty-five participants completed the trial, while five discontinued prematurely due to: two or more relapses (n = 2), three or more courses of steroid pulse therapy given (n = 1) or lymphocyte counts in the pretreatment sample failing to meet screening criteria (n = 2); discontinuations were evenly distributed between groups (**Figure 1A&B**, **Supplementary Table 3**). One serious adverse event, a Grade 3 gastroenteritis, occurred in the placebo group and resolved promptly without trial discontinuation (**Supplementary Table 4**). No significant differences in safety profiles emerged between OCH and placebo, confirming that OCH was well tolerated (**Supplementary Figure 1**, **Supplementary Table 5**).

**Figure 1.**
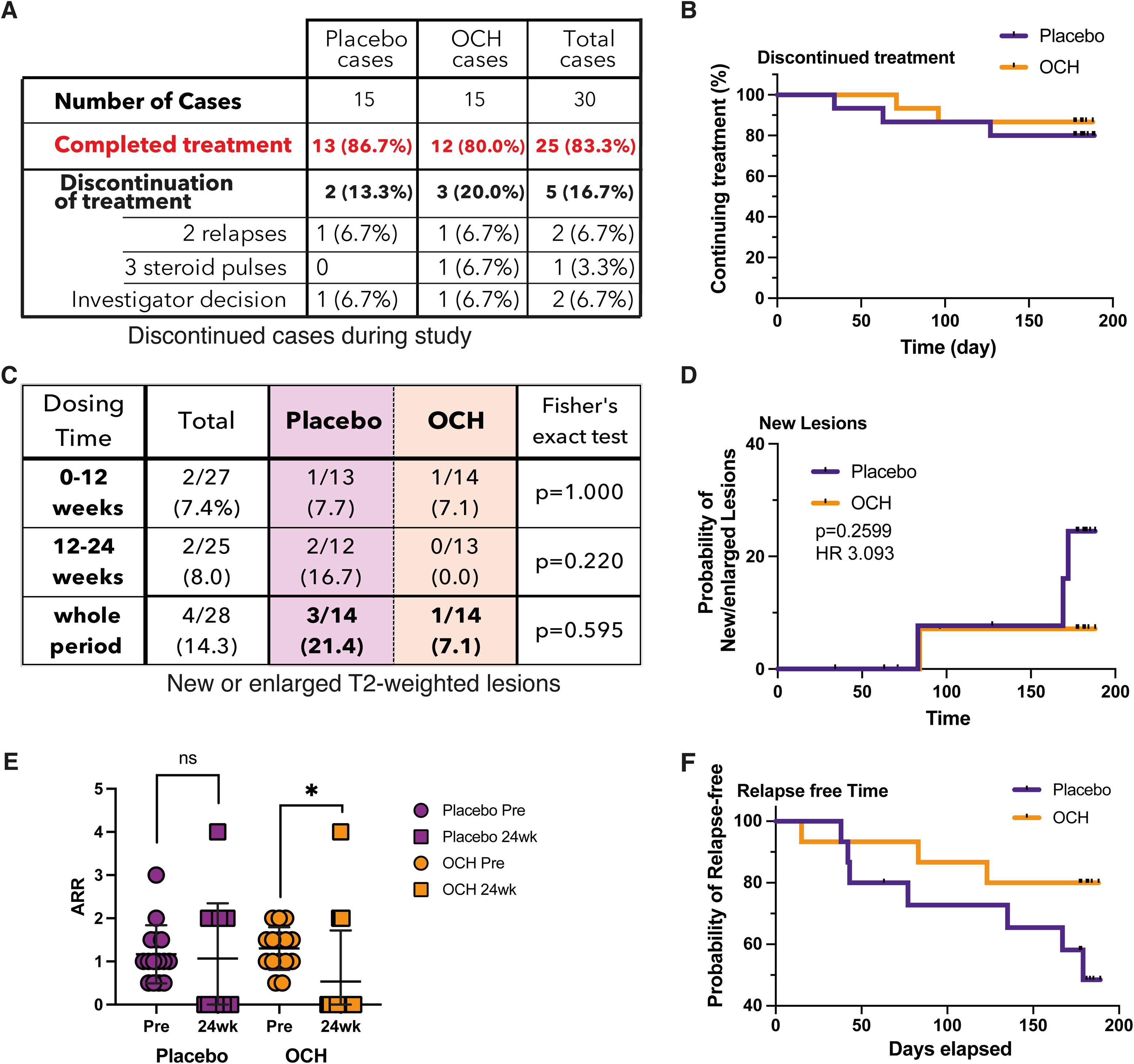
OCH treatment was associated with fewer new lesions and reduced relapses. (A) Summary of patient categories and reasons for treatment discontinuation during the study period. Detailed information is provided in Supplementary Table 3. (B) Kaplan-Meier (KM) curves of continuation in the trial for all participants, with time to discontinuation indicated for the five discontinued cases. (C) Lesions were assessed at baseline and at 3 and 6 months during the trial by expert radiologists using gadolinium-enhanced MRI. The number of patients with newly-developed lesions at the end of the trial is summarized by treatment group. (D) Reverse KM curves of time to first new lesion, divided by treatment group (Placebo, n = 14; OCH, n = 14; Mantel-Cox Log-Rank test). (E) Annualized relapse rates (ARR) calculated for each patient during the 2 years preceding trial treatment initiation (Pre) and compared with the ARR during the treatment period (24 weeks). Data are shown by treatment group (Placebo, n = 14; OCH, n = 14; mean ± sd; paired t-test). (F) KM curves of relapse-free survival among all patients with relapsing MS divided by treatment group (Placebo, n = 14; OCH, n = 14; Mantel-Cox log-rank test). For panels C-F, one placebo-treated and one OCH-treated patient who failed to meet screening criteria because of low lymphocyte counts were excluded from the analysis. Treatment was discontinued at day 63 in the placebo group and day 71 in the OCH group. No new lesions or relapses were observed in these two patients during the limited observation period.

### OCH prevents relapse in patients with SPMS, but not RRMS

The primary trial outcome measure, new or enlarged T2-weighted MRI lesions, was not statistically met, given the low new lesion counts across all groups. Four patients developed new lesions, with fewer observed in the OCH group (**Figure 1C&D; Supplementary Table 6)**. Lesion number was not correlated with baseline counts, and lesion volume remained stable in both groups (**Supplementary Figure 2A&B**). New lesions clustered towards the end of the study period, suggesting this was too short duration for this measure (**Figure 1D; Supplementary Figure 2C**).

Analysis of secondary outcomes revealed OCH strikingly reduced the annualized relapse rate (ARR) compared with pre-enrolment rates (**Figure 1E&F, Supplementary Table 7**). Relapse-free survival was substantially extended in OCH-treated SPMS, but not RRMS (**Figure 2**). Remarkably, all 6 OCH-treated SPMS patients remained relapse-free, while 5 out of 6 placebo-treated SPMS patients experienced relapses. These data indicate a clinically meaningful benefit of OCH in SPMS and support its potential as a therapeutic option.

**Figure 2.**
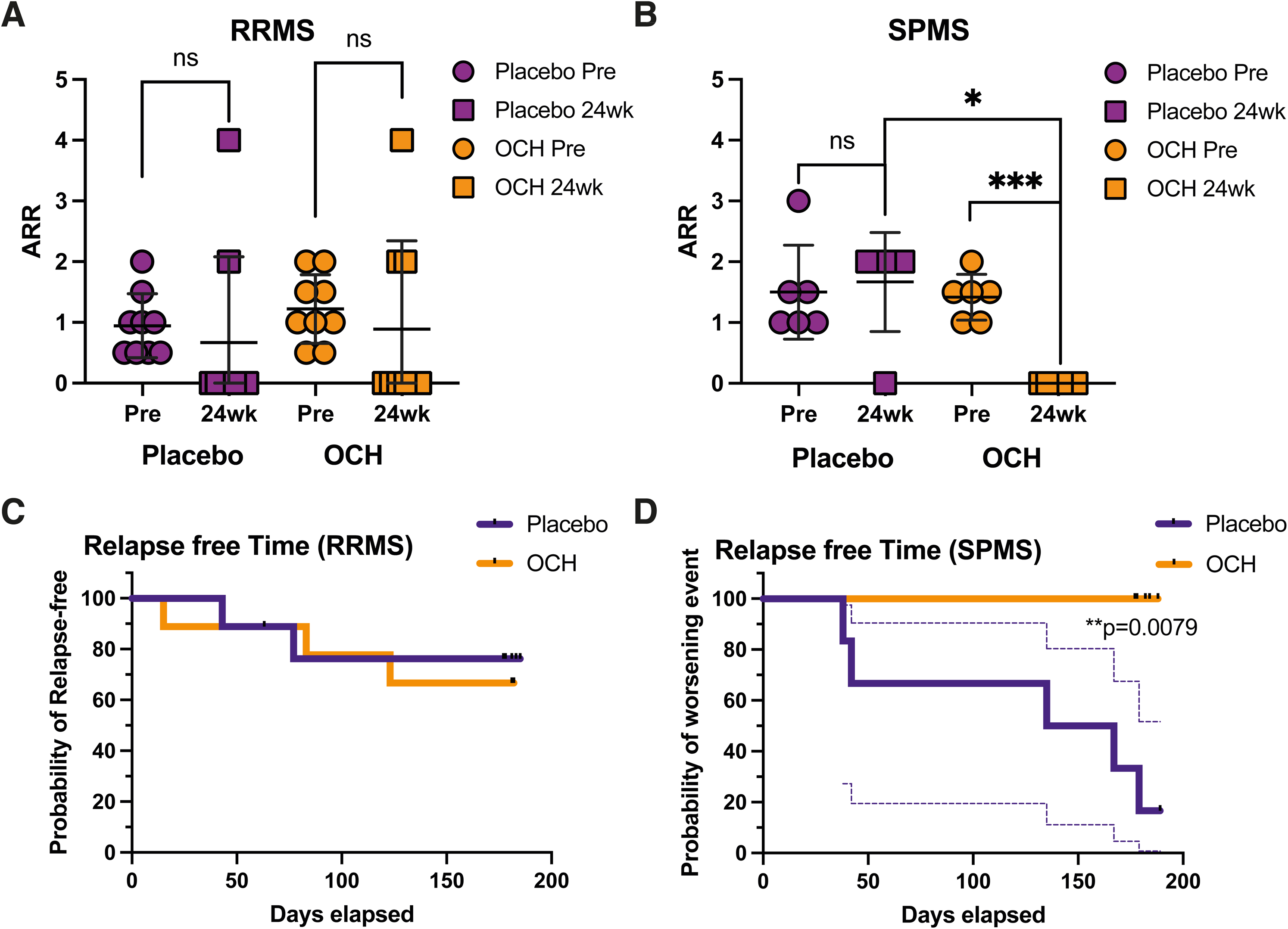
Effects of OCH on relapse suppression. (A) ARR for placebo and OCH-treated patients with RRMS at pre-and 24 weeks after the treatment period (Placebo, n = 8; OCH, n = 8); mean ± sd; paired *t*-test. (B) ARR for placebo and OCH-treated patients with SPMS at pre- and 24 weeks after the treatment period (Placebo, n = 6; OCH, n = 6); mean ± sd; paired *t*-test. (C) KM curves of relapse-free survival in placebo and OCH-treated RRMS patients (Placebo, n = 8; OCH, n = 8). (D) KM curves of relapse-free survival in placebo and OCH-treated SPMS patients (Placebo, n = 6; OCH, n = 6). 1 placebo-treated and 1 OCH-treated patient who failed to meet screening criteria at first study visit were excluded as noted.

### OCH-treated patients have reduced risk of disease activity

PIRA, defined as 12 to 24-week confirmed disability progression excluding RAW (*22*), can be measured by Expanded Disability Status Scale score changes (ΔEDSS) or functional scale scores changes (ΔFS) in a period where there were no relapses. Over the study period, worsening of EDSS was observed in only a single patient, while a few patients had worsening or improvements in FS (positive and negative ΔFS), overall changes in measures of PIRA were not significant between groups (**Supplementary Figure 3A-D**). Brain volume changes, assessed by MRI, also did not differ between groups (**Supplementary Figure 3E**).

By integrating outcome measures, patients experiencing disease worsening could be directly classified separately to those patients who showed no evidence of disease activity (NEDA-3; criteria: no new or enlarging MRI lesions, no clinical relapse, and no evidence of disability progression or PIRA (*23–25*)). The classification of NEDA versus worsening (non-NEDA) is depicted for individual cases (RRMS: **Figure 3A**; SPMS: **Figure 3B**) and summarized for the overall groups in **Supplementary Table 8**. Among RRMS patients, 4/8 receiving placebo and 5/8 treated with OCH achieved NEDA-3 over the 6-month study period. For SPMS, 5 of 6 OCH-treated patients maintained NEDA, whereas no placebo-treated SPMS patients maintained NEDA.

**Figure 3.**
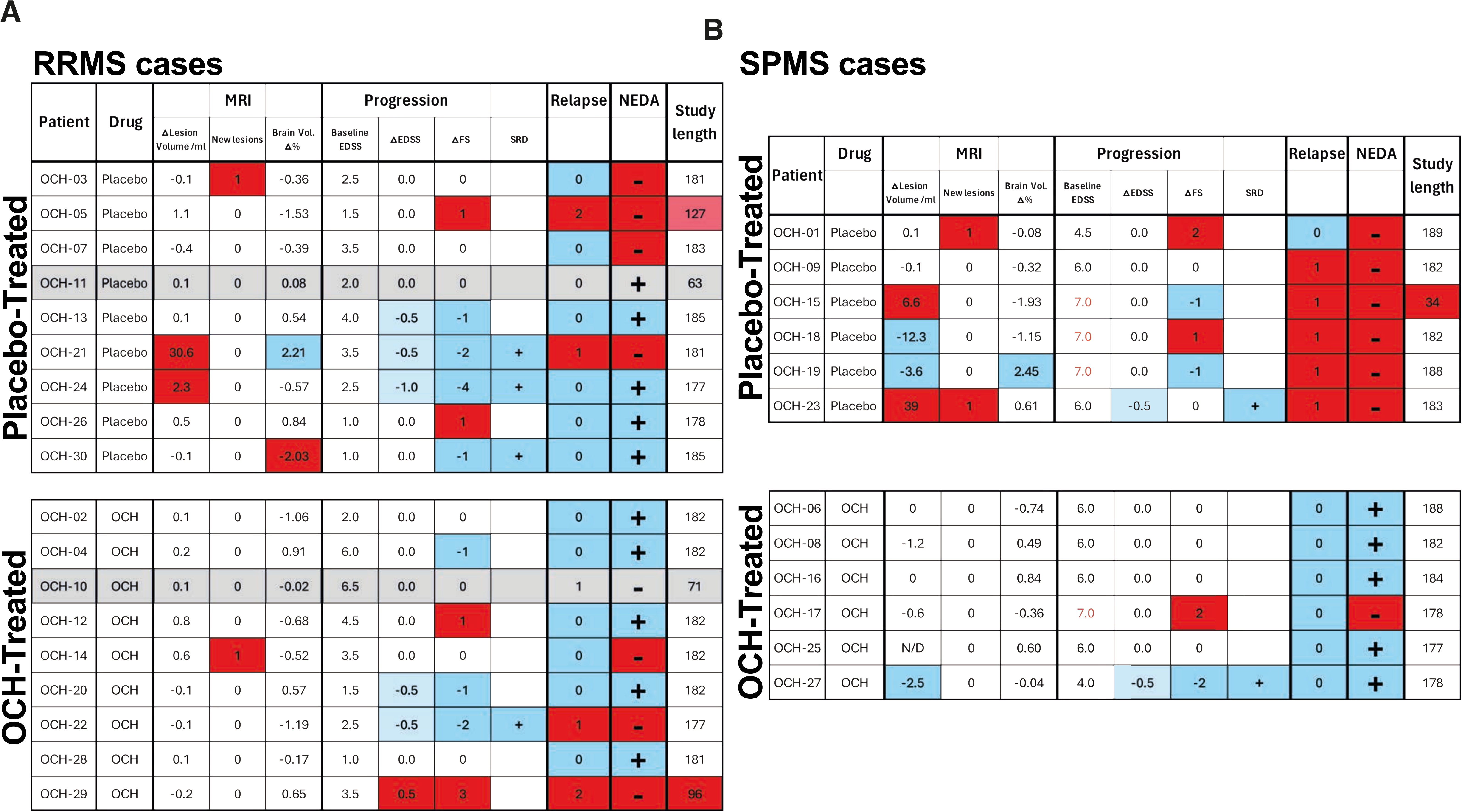
Comparison of OCH and placebo treatment effects in individual cases. Changes in clinical and radiological observations in individual patients during the trial are summarized separately for RRMS (A) and SPMS (B) and divided by treatment group (placebo, upper tables; OCH, lower tables). Changes observed during the study period are highlighted in red for worsening and in blue for improvement according to the following criteria: change in total lesion volume (Δlesion volume ≥ 2 ml); occurrence of new lesions during the study period (>0); percentage change in brain volume (negative values indicate atrophy, thresholds +/-2%); change in EDSS score (ΔEDSS, any positive change indicates increased disability, negative change indicates reduced disability); change in FS score (ΔFS, any positive change indicates increased disability, negative change indicates reduced disability); sustained reduction in disability (SRD, any confirmed reduction is an improvement); relapse occurrence during the study period (any relapse vs. no relapse); and NEDA status (any disease activity observed vs. NEDA-3 achieved over the study period). Study duration is also indicated, including cases in which treatment was discontinued because of disease recurrence or failure to meet screening criteria.

The sole worsening OCH-treated SPMS case had a very high level of disability at baseline and showed only a slight increase in upper limb impairment, as measured by FS scoring. In contrast, worsening in placebo-treated SPMS patients included new MRI lesions, increased disability scores, and clinical relapses.

Overall, OCH-treated cases trended toward a longer time to disease worsening and more likely to maintain NEDA status during the trial (p = 0.0570; OR = 6.25; **Figure 4A&B**). Divided by disease sub-type, OCH protected SPMS patients from disease worsening, significantly extending the time to disease worsening (p = 0.0049; **Figure 4C**) and achieving NEDA-3 in most cases (p = 0.0152, OCH vs placebo; **Figure 4D**); whereas RRMS only saw a modest boost in sustained NEDA under OCH treatment (OR = 1.67).

**Figure 4.**
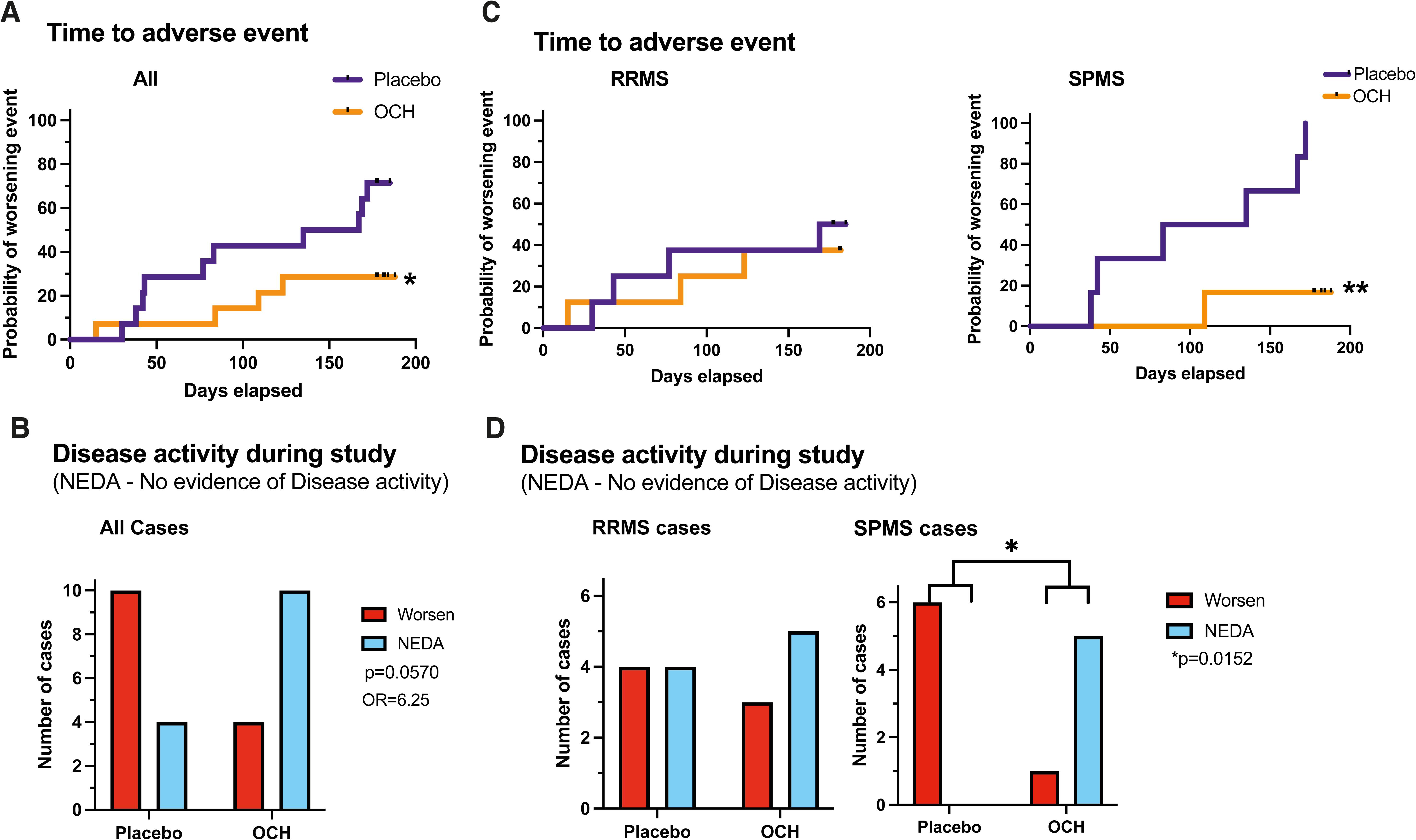
OCH-treated relapsing MS patients maintain NEDA. (A) KM curves for time to disease worsening defined clinically/radiologically (new lesions, relapse, increase in EDSS or FS scores) divided by placebo or OCH treatment. (B) Comparison of number of patients maintaining NEDA-3 (NEDA, blue) versus cases with 1 or more worsening measure (new lesion, relapse, increase in EDSS or FS; Worsen, red) over the study period divided by treatment group for all patients. (C) KM curves for time to disease worsening separated into RRMS (left), and SPMS (right). (D) Contingency analyses of NEDA-3 (blue) vs worsening cases (red) for RRMS (left), and SPMS (right). Statistical significance of KM curves was assessed using a Mantel-Cox Log-Rank test and contingency testing was by Fisher’s exact test between placebo and OCH-treated; all cases (Placebo, n = 14; OCH, n = 14), RRMS (Placebo, n = 8; OCH, n = 8); SPMS (Placebo, n = 6; OCH, n = 6); the 2 RRMS cases discontinued for screening criteria were excluded from these analyses.

### Alterations of iNKT cells and IL-4-producing Th cells in OCH treated patients

To explore the cellular mechanisms underlying the therapeutic effects of OCH on relapsing MS (rMS), we performed flow-cytometric analysis of peripheral blood lymphocyte populations from patients treated with OCH or placebo. As OCH targets iNKT cells, we first focused on these cells. However, peripheral blood contains only low numbers of iNKT cells(*14*) with even lower number in MS patients(*26, 27*). Total iNKT cell numbers were unchanged after OCH treatment (**Figure 5A**), yet CD4^+^ iNKT cells rose significantly in OCH-treated patients compared with baseline (p=0.0342; **Figure 5B&C**). Interestingly, CD4^+^ iNKT cells are suggested as an immunoregulatory type associated with IL-4 production that increases during MS remission, whereas CD4^-^ iNKT cells are associated with inflammatory cytokines(*27, 28*).

**Figure 5.**
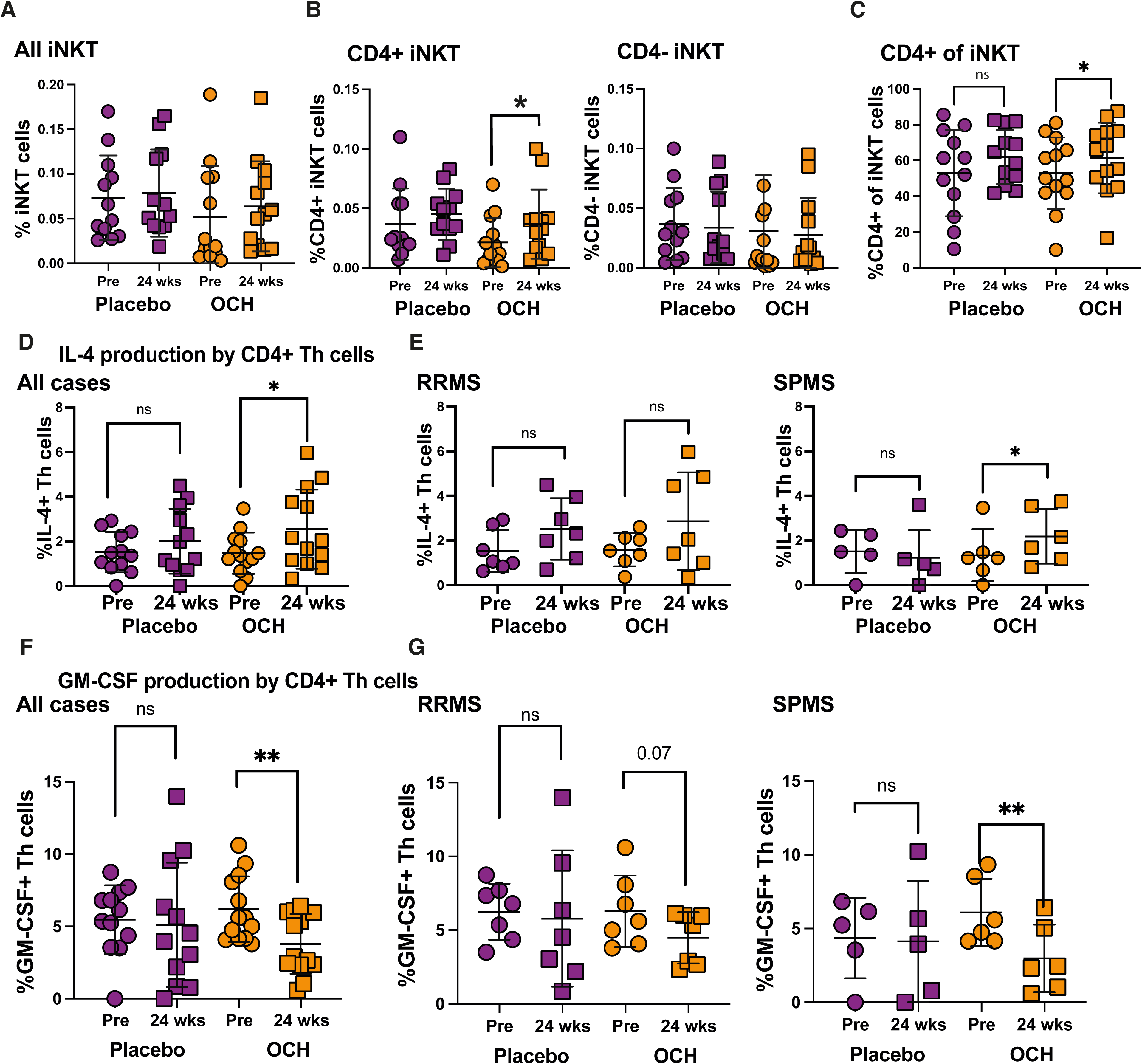
OCH increases IL-4+ Th cells in decreases GM-CSF+ Th cells. Peripheral blood was collected at baseline prior to first drug administration and at week 24. Peripheral blood mononuclear cells (PBMCs) were isolated and immune cell subsets were assessed by flow cytometry. iNKT cells were defined as CD3⁺CD56⁺PBS-57/CD1d tetramer+ cells and Th cells were defined as CD3⁺CD4⁺CD56⁻. (A) Proportions of total cells within PBMCs for all iNKT cells. (B) Proportions of CD4⁺ iNKT cells (left), and CD4⁻ iNKT cells (right). (C) Proportion of CD4-expressing cells within the total iNKT cell population. Pre/post treatment values were compared using the Wilcoxon matched-pairs signed-rank test. PBMCs were restimulated with PMA/ionomycin and cytokine-producing Th cells were measured. (D) Comparison of levels of IL-4+ Th cells between baseline and week 24 for placebo and OCH-treated groups for all patients. (E) Comparison of levels of IL-4+ Th cells between baseline and week 24 for placebo and OCH-treated groups for RRMS (left), and SPMS (right). (F) Comparison of levels of GM-CSF+ Th cells between baseline and week 24 for placebo and OCH-treated groups for all patients. (G) Comparison of levels of GM-CSF+ Th cells between baseline and week 24 for placebo and OCH-treated groups for RRMS (left), and SPMS (right). Plots show mean ± sd and data were tested with paired *t*-tests; *p<0.5, **p<0.01; patients who discontinued the study were excluded; placebo: RRMS n = 7, SPMS n = 5; OCH: RRMS n = 7, SPMS n = 6.

Our previous studies demonstrated OCH selectively induced IL-4 production by iNKT cells over the prototypic iNKT cell ligand α-GalCer (*15, 16*). As IL-4 canonically drives Th2 differentiation and OCH treatment shifted cells towards Th2 in EAE (*16*), we examined IL-4 producing Th cells in this study. In comparison with placebo-treated rMS, OCH-treated rMS group showed a significant increase in IL-4-producing Th cells in the peripheral blood after 24 weeks (p=0.0310). Similar results were obtained when we focused on the SPMS group (p=0.0338) (**Figure 5D&E**).

### OCH treatment decreases circulating GM-CSF^+^ Th cells

As OCH treatment led to increased IL-4-producing Th cells, we also evaluated the effects of oral OCH treatment on circulating CD4^+^ T cells that are potentially pathogenic in MS, including Th1, Th17 and GM-CSF-producing Th cells (*29, 30*). IL-17 and IFN-γ-producers were unchanged by placebo or OCH treatment (**Supplementary Figure 4A&B**). However, GM-CSF-producing Th cells, which have recently been linked to pathogenicity in MS (*31–33*), decreased significantly under OCH treatment at 24 weeks compared with baseline (p=0.0022) (**Figure 5F**). Decreases in GM-CSF-producing Th cells occurred in both RRMS and SPMS groups, with reductions most apparent in SPMS (**Figure 5G**) and in cases achieving NEDA status (**Supplementary Figure 4C**).

### OCH alters pathogenic Th cells in mouse models of MS

To advance the project within a translational research framework, we conducted mechanistic studies using an animal model of MS. OCH treatment ameliorated standard EAE (**Figure 6A**), a preclinical model for MS suggested as representative of an acute relapse(*34*). OCH treatment did not significantly affect CNS-infiltrating IFN-γ^+^ and IL-17^+^ Th cells (**Supplementary Fig 5A)**. This is intriguing considering the strong decrease in disease as Th1 and Th17 are considered to be the major pathogenic Th subsets in EAE(*35*). GM-CSF-producing Th cells, a pathogenic population that is critical determinant of EAE disease induction that can arise from either Th1 and Th17 lineages(*36, 37*), were significantly reduced in the CNS of OCH-treated mice compared with controls (**Figure 6B**). *Ex vivo* stimulation of CNS-infiltrating T cells with the immunising peptide showed lower secretion of IL-17, IFN-γ, and GM-CSF by cells derived from OCH-treated mice versus those from control mice (**Supplementary Figure 5B**).

**Figure 6.**
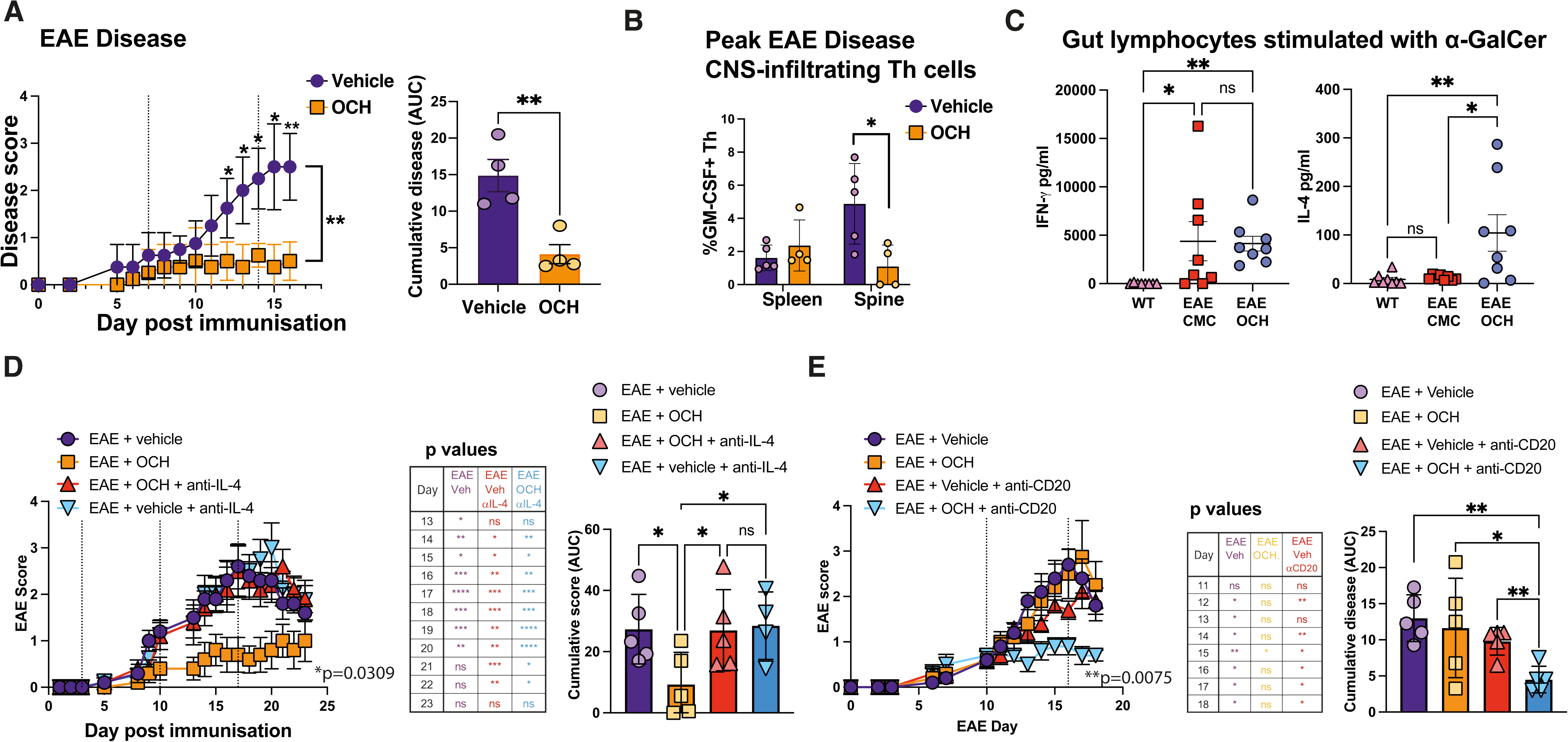
OCH modulates Th cells and disease in EAE in an IL-4-dependant manner. EAE was induced in mice by MOG_35-55_ immunization with groups of mice receiving oral OCH or vehicle (CMC) at Day 0 and weekly intervals thereafter as noted by dotted lines. (A) Daily EAE scores (left) and cumulative disease (area under the curve, right). Data are representative of at least 6 similar independent experiments. (B) At peak EAE, spleen or spinal-infiltrating Th cells (CD45+CD11b-TcRβ+CD4+CD8-) were isolated, restimulated with PMA/ionomycin, and assessed by intracellular flow cytometry for GM-CSF-producing Th cells. n = 4-5 per group, data are representative of 2-4 independent experiments. (C) C56BL/6 mice were immunized to induce EAE and were treated orally with either OCH or vehicle (CMS). On day 7, small intestine lamina propria lymphocytes were isolated and restimulated with α-GalCer. Wild type littermate mice (WT) were examined similarly as a control. After 48 hours, cytokine concentration in supernatants was assessed by ELISA for IFN-γ (left) or IL-4 (right). WT, n=7; EAE/CMC, n=8; EAE/OCH, n=8. Data are representative of 2 independent experiments. (D) Groups of C57BL/6 mice in which EAE had been induced were treated with either oral OCH or CMC (vehicle) on Days 3,10,17 and with either anti-IL-4 depleting or isotype control antibodies on day 4 and 8. EAE was scored daily (left) and cumulative disease was calculated from area under the curve (right). *p* values from daily comparisons between EAE/OCH and other groups are shown in the center table. n=5 per group; data are representative of 2 independent experiments. (E) Groups of C57BL/6 mice in which EAE had been induced were treated with either oral OCH or CMC (vehicle) on day 10 and 16 and with either anti-CD20 depleting or isotype control antibodies on day -6 and 1. EAE was scored daily (left) and cumulative disease was calculated from area under the curve (right). *p* values from daily comparisons between OCH/anti-CD20 and other groups are shown in the center table. n=5 per group; data are representative of 4 similar experiments. For EAE studies: EAE scores were tested by two-way ANOVA with Bonferroni-corrected multiple comparison for individual points; cumulative disease scores were tested with Mann-Whitney U tests; cell populations were tested with unpaired t-tests with Welch’s correction where required.

### OCH mediates its effects by regulation of IL-4

OCH is a CD1d-dependent activator ligand for iNKT cells, thus the observed increases in Th2 cells likely stem from indirect effects from IL-4 elaborated by activated iNKT cells. Both EAE experiments and our human phase I trial (*19*), indicated that immunological effects of orally administered OCH can be detected in peripheral blood as early as 2 hours after administration. This rapid response is consistent with the notion that oral OCH activates iNKT cells directly within the gut mucosa. Indeed, stimulating intestinal mucosa-derived lymphocytes with the specific iNKT-cell agonist α-GalCer showed gut iNKT cells from mice treated with OCH had specifically increased IL-4 production (**Figure 6C**). These data support the concept of oral OCH leading to prior *in vivo* activation and long-lasting polarization of gut-resident iNKT cells towards a Th2-inducing phenotype.

Furthermore, we showed that suppression of mouse EAE by oral OCH was largely abrogated by administration of neutralizing anti-IL-4 antibodies (**Figure 6D**), confirming IL-4-dependence as previously indicated(*16*). These findings underscore iNKT-mediated disruption of pathogenic Th cell development via IL-4 in OCH-mediated immunomodulation.

### OCH modulates B cell activation affecting efficacy

Although OCH administration was associated with reduction in GM-CSF-producing pathogenic Th cells in both RRMS and SPMS, it remains unclear why OCH was predominantly effective in SPMS patients. Some differences previously highlighted between pathogenesis in RRMS and SPMS include alterations in the location and functional properties of B cells (*38–40*) and B cell-depleting therapies show a more limited benefit in SPMS compared with RRMS(*41–43*). It is conceivable, in addition to modulating pathogenic Th cells, that iNKT cell-produced IL-4 could alter B cell activation. In support of this hypothesis, OCH treatment amplified EAE-associated increases in Class-switched B cells, germinal center and activated B cells (**Supplementary Figure 5D**), and increased serum IgG (**Supplementary Figure 5E**). Similarly, OCH treatment of RRMS patients led to increased serum levels of IgG1, IgG2, and IgG3, whereas IgG4, IgA, and IgM were not altered significantly (**Supplementary Figure 6**). These data suggest that OCH may stimulate B cell activity indirectly as well as modulating Th cell pathogenicity.

When mice were treated with a delayed, suboptimal OCH treatment regimen, only limited effects on clinical disease signs were observed in conventional EAE; in contrast, this OCH treatment ameliorated clinical EAE in anti-CD20 B cell-depleted mice (**Figure 6E**). The enhancement of responsiveness to OCH by B cell depletion may explain the selective efficacy in SPMS, with augmentation of B cell mediated pathogenesis in RRMS counterbalancing suppression of pathogenic Th cells. Thus, combination therapy in which pro-inflammatory B cell activity is adequately suppressed at the same time as OCH acts on T cells may provide greater efficacy in RRMS. While this hypothesis requires further investigation, it raises the possibility that interactions between B cells and pathogenic Th cell responses may contribute to the differential efficacy of OCH across MS subtypes.

## DISCUSSION

This phase II trial demonstrates that the synthetic glycolipid OCH reduces relapses in MS, with unprecedented efficacy in SPMS, a disease course historically refractory to immunomodulation. OCH prevented clinical worsening, achieving NEDA-3 in most patients with a tendency towards fewer new lesions in SPMS. Whilst highly effective agents are now available for RRMS, their impact on SPMS remains limited, rendering this chronic disease stage an unmet and increasingly important clinical need(*39*). Unlike existing therapies, OCH achieves these benefits without depleting major immune cell populations. Instead, it reprograms pathogenic T cell responses through IL-4–biased activation of iNKT cells. These findings identify a gut-directed, non-depleting immunomodulatory pathway capable of regulating CNS autoimmunity and support a new therapeutic paradigm for progressive disease.

iNKT cells are rare in peripheral blood but enriched in tissues such as the liver and gut, (*44*) where they recognize glycolipid antigens derived from bacteria (*45, 46*) and act as sensors of the gut-environment regulating systemic immunity. A role for iNKT cells in the relationship between gut and autoimmunity is highlighted as iNKT cells are critical for EAE amelioration by antibiotic treatment (*47*). In preclinical models, activation of iNKT cells was initially shown to ameliorate autoimmune disease, including EAE and diabetes(*48, 49*), establishing this pathway as a compelling therapeutic target. However, subsequent studies revealed that the prototype ligand α-GalCer could exert bidirectional effects: depending on dose, timing, route, and disease context, α-GalCer not only suppressed disease but could also exacerbate autoimmunity by enhancing pathogenic Th1 or Th17 responses (*50, 51*). This intrinsic instability and dose sensitivity ultimately dampened enthusiasm for iNKT-directed therapies for autoimmune disease(*52*). Subsequent in-human studies have been focused on productive iNKT activation for cancer immunotherapy via strengthening Th1 responses (*53, 54*).

Not all iNKT ligands are equivalent, we showed that the α-GalCer analogue OCH possesses a distinct and highly advantageous immunological profile. Although OCH retains the same TcR contact region as α-GalCer, its truncated sphingosine chain alters CD1d:TcR interaction kinetics, resulting in shorter dwell times and a qualitatively different signalling outcome(*15, 55*). This briefer engagement preferentially induces early IL-4 transcription while limiting the sustained signalling required for IFN-γ production. Functionally, OCH elicits a robust and reproducible IL-4-dominant response from iNKT cells without the unpredictable dose-dependant characteristic of α-GalCer. In multiple preclinical models, including EAE, arthritis, diabetes, and colitis, OCH consistently ameliorated disease (*16–18, 56*). These properties uniquely position OCH as a ligand capable of harnessing iNKT cells for immune regulation in a predictable and therapeutically tractable manner.

As a once-weekly oral agent, OCH offers a mechanistically coherent target, directly engaging mucosal immunity. Our data demonstrate a selective effect on gut-resident iNKT cells, providing a plausible explanation for its rapid systemic immunological impact and aligning with the emerging paradigm that the gut–brain axis and microbiome are central to MS pathogenesis(*57*). NKT cells are reduced in MS, potentially reflecting gut dysbiosis (*26, 27, 58*). Interestingly, oral (*59*)OCH increased regulatory type CD4⁺ iNKT cells(*60*), which are associated with IL-4production in human(*59*). As the intestine is increasingly recognised as a major site of pathogenic Th cell education(*37*) and gut dysbiosis in MS is associated with increased Th17 cells(*61*), targeting this compartment represents a strategically attractive means of reprogramming disease-driving immune networks.

OCH treatment was associated with a reduced frequency of GM-CSF-producing Th cells in both EAE models and MS, a Th cell subset type implicated in MS pathogenesis (*31, 33*). Intestinal Th17 cells are thought to give rise to pathogenic GM-CSF–producing Th cells following IL-23–dependent stimulation, (*37*)and IL-4 is a potent antagonist of IL-23 signalling (*62*). It is therefore plausible that OCH, through induction of IL-4 by mucosal iNKT cells, interferes with the emergence of this pathogenic lineage at an early differentiation stage. This interpretation remains speculative, but provides a plausible mechanistic link between gut iNKT cell activation, IL-4 production, and the suppression of pathogenic Th cells. Therefore, orally-administered OCH may specifically act within the gut to reprogramme Th cell fate, favouring Th2 responses whilst limiting generation of pathogenic effector cells.

Oral OCH was previously shown safe and immunomodulatory in phase I, with rapid effects on peripheral immune cell populations (*19*). In this phase II trial, OCH reduced disease activity in relapsing MS, showing trends towards fewer lesions formation. In SPMS, a disease stage historically refractory to treatment, OCH achieved high NEDA-3 rates. Critically, no OCH-treated SPMS suffered relapses or new lesions; in contrast to relapses in more than 80% of placebo-treated SPMS patients with new lesions in 30%.

The primary endpoint, new/enlarging T2 lesions, was not met in this study; reflecting low event rate over the short 6-month treatment period (4 new lesions across all 30 cases). This should be interpreted in the context of well-established kinetic dissociation between immunological efficacy and MRI readouts(*63*), particularly for therapies that modulate rather than deplete immune responses. For example, glatiramer acetate, which also rebalances T cell activation, has a delayed effect on MRI activity despite clear long-term clinical benefit(*64*). Even for highly potent depleting therapies such as Ocrelizumab (anti-CD20), showed evidence of persistent active inflammation by MRI in first 6 months of treatment; with new T2 lesions occurring almost exclusively in the first 24 weeks (*65*) and robust reduction of lesions observed only after 12-24 months. These observations likely reflect biological inertia within the CNS, including the persistence of resident pathogenic immune cells and ongoing innate activation despite rapid changes in the periphery. Thus, disease activity detected by MRI early in the treatment does not necessarily imply therapeutic failure, but rather the lag required for compartmentalised inflammation to resolve. This issue is particularly apparent in SPMS, where trials typically require large cohorts and extended follow-up to detect modest effects, and where siponimod remains the only approved therapy with limited impact on progression(*8*). Against this backdrop, the consistent clinical protection observed with OCH, despite this being a small-scale study, is exciting and valuable. Although this study was not powered to detect radiological change, the absence of relapses and new lesions in OCH-treated SPMS, contrasted with frequent activity in placebo-treated patient, highlights potential biological and clinical importance that merits confirmation in larger and longer studies.

Although OCH induced clear immunological shifts in RRMS, including increased IL-4-producing T cells and reduced GM-CSF⁺ Th cells, this did not translate into a comparable clinical benefit. One explanation is that IL-4 exerts divergent effects in RRMS: while it constrains pathogenic Th differentiation, it is also a potent enhancer of B cell activation, antibody production, and antigen presentation. Given the central role of B cells in RRMS pathogenesis, as underscored by the efficacy of B cell-depleting therapies(*10, 11*), iNKT-derived IL-4 augmenting B cell activity (*66*) could counterbalance the beneficial effects of OCH at this disease stage. Supporting this interpretation, we observed in EAE that the therapeutic impact of OCH was markedly enhanced by experimental B cell depletion, indicating that B cells modulate the net clinical effect of iNKT-directed therapy. In contrast, peripheral B cells appear to play a more limited role in SPMS, where pathology is increasingly compartmentalised within the CNS(*67–69*). Under these conditions, IL-4-mediated rebalancing of Th networks may exert unopposed benefits, similar to the striking efficacy observed in SPMS.

These findings suggest that OCH may be intrinsically active in both RRMS and SPMS, but that its clinical impact in RRMS is constrained by concurrent B cell–driven pathology. This raises the rational possibility that combining OCH with B cell-targeted therapies could unmask its full therapeutic potential in RRMS, while preserving its unique capacity to modulate pathogenic T cell networks.

Despite its short duration and small cohorts, this Phase II trial demonstrates that oral OCH is safe, biologically active, and clinically meaningful in relapsing MS, with a particularly striking effect in SPMS. Relapses in SPMS are now recognized as a major contributor to irreversible disability, as recovery in this disease stage is typically incomplete. The prevention of relapse activity in SPMS therefore represents a clinically important therapeutic advance.

OCH defines a new class of immunomodulatory therapy in MS: a non-depleting, orally administered agent that acts through the gut-iNKT/IL-4 axis to rebalance pathogenic T cell networks. These findings challenge the prevailing view that immune modulation is largely ineffective in progressive disease and support the concept that targeted immune reprogramming remains viable in SPMS. Together, these data provide a strong rationale for larger and longer trials of OCH in SPMS, and for mechanistically informed studies in RRMS, including rational combination strategies. By linking a defined immunological pathway to a clinically meaningful signal in a historically treatment-refractory population, this study establishes both a therapeutic opportunity and a framework for stage-specific intervention in multiple sclerosis.

## Materials and Methods

### Study design and participants

This study was a double-blind, multi-center, placebo-controlled, randomized, phase II clinical trial (www.clinicaltrials.gov registration number NCT04211740). Thirty patients with relapsing MS who met the study criteria were recruited from three hospitals (NCNP, Tokyo Medical and Dental University Hospital and Tokyo Women’s Medical University Hospital) between September 2019 and June 2021. The study design has been reported previously (*70*) and inclusion criteria are also summarised in **Supplementary Table 1**. Patients were randomly selected to receive placebo or OCH using a clinical-based management system (eClinical Base®, Translational Research Center for Medical Innovation, Kobe, Japan) to ensure balanced allocation. Participants took 300 mg granules orally with water once per week for 24 weeks. The granules were composed of crystalline cellulose, mannitol, sodium croscarmellose, low-substituted hydroxypropyl cellulose, and polysorbate 80. The OCH formulation additionally contained 3.0 mg OCH and these granules and packaging were identical to the placebo granules except for the active compound. Both participants and investigators were blinded to treatment allocation . The study protocol was approved by the institutional ethics committees of the participating institutions and all participants provided written informed consent.

The primary outcome measure was subjects with new or enlarged lesion at 24 weeks detected by T2-weighted MRI. The secondary outcomes included annual relapse rate (ARR), relapse-free period, duration of no evidence of disease activity (NEDA), sustained reduction in disability (SRD) occurrence, and exploratory biomarkers from Phase I trials (*19*), including immune cell subsets in peripheral blood.

### Patient assessment

Patients received first dose as in-patients and were assessed for adverse effects on the following day. Thereafter, participants attended outpatient clinics once per month for medical examination. Neurologic assessments were performed at baseline and at monthly intervals by a blinded expert neurologist and included documentation of clinical exacerbations and current disability severity; scoring criteria for Enhanced Disability Status scale (EDSS) and Functional System (FS) have been modified from the original scales (*71, 72*) to align with the Japanese medical system as defined by the Japanese Intractable Diseases Information Center (https://www.nanbyou.or.jp/).

### Magnetic resonance imaging analysis

Brain magnetic resonance imaging was performed in all participants using a 3-T MR system (Philips, Best, The Netherlands or Siemens, Munich, Germany) with gadolinium enhancement. MRI scans were obtained within 1 week prior to initiation of OCH administration and repeated either 90 days later or at the time of treatment discontinuation. The MRI analyses included: number of new lesions on T2-weighted MRI images; number of new low-signal foci on T1-weighted MRI images; and number of MS lesions on gadolinium contrast-enhanced T1-weighted MRI images. MRI imaging was assessed by an expert neuroradiologist who was blinded to the study. Brain volumes and lesion volumes were measured on 3D-T1W1 using FreeSurfer 6.0 neuroimaging analysis software (Athinoula A. Martinohes Center for Biomedical Imaging, Harvard Medical School, MA, USA) (*73*) or by the lesion growth algorithm (*74*) on T2/3D-FLAIR using the LST toolbox 3.0.0 for SPM.

### Flow cytometric assessment of patient immune cells

Peripheral blood samples were collected from participants at baseline (prior to treatment), and at 1 day and 24 weeks after initiation of treatment. Peripheral blood mononuclear cells (PBMCs) were isolated by Ficoll-Paque density-gradient centrifugation (Ficoll-Paque Plus, GE Healthcare, Marlborough, MA, USA) and resuspended in PBS supplemented with 5% foetal calf serum.

Leukocyte subset assessment: PBMC were stained with fluorochrome-conjugated antibodies against surface antigens. Flow cytometry antibodies were purchased from BD Biosciences (Franklin Lakes, NJ, USA), Biolegend (San Diego, CA, USA), Beckman Coulter (Indianapolis, IN, USA), or eBioscience brand (Thermo-Fisher, Waltham, MA, USA) and a list of antibodies used in this study is provided in **Supplementary Table 9**.

Th cell subset assessment: PBMC were stimulated with PMA (50 ng/ml) and ionomycin (1000 ng/ml) (both Sigma-Aldrich, St. Louis, MO, USA) in the presence of Golgi Stop (BD Biosciences) for 4 h in AIM V media (Thermo) at 37°C, 5% CO_2_ in a humidified incubator. Cells were surface stained as above before fixation with eBioscinece Transcription Factor Staining kit (Thermo) according to manufacturer’s instructions. Fixed cells were then stained for fluorochrome-conjugated antibodies against intracellular antigens.

Flow cytometry data were acquired using a FACSCanto II with BD FACSDiva 5.0 software (BD Biosciences) and analyzed using FlowJo software version 10.10.0 (BD Biosciences).

### Animals, EAE induction and treatment

For acute EAE, C57BL/6 mice were purchased from CLEA Japan, Inc. (Tokyo, Japan). Mice aged 6-8 weeks were used in experiments and were maintained in specific pathogen-free conditions and all animal experiments were approved by the Committee for Small Animal Research and Animal Welfare of the National Center of Neurology and Psychiatry. All efforts were made to minimize animal suffering in experiments.

EAE induction was carried out as previously described (*75*). Briefly, mice were injected subcutaneously with a MOG_35-55_ peptide (100 μg; synthesized by Toray Research Center, Tokyo, Japan) and heat-killed Mycobacterium tuberculosis H37RA emulsified in complete Freund’s adjuvant (1 mg; Difco, KS, USA). Pertussis toxin (100 ng; List Biological Laboratories, CA, USA) was injected intraperitoneally on days 0 and 2 after immunization. EAE was clinically scored daily (0, no clinical signs; 0.5, tail weakness; 1, partial tail paralysis; 1.5, severe tail paralysis; 2, flaccid tail; 2.5, flaccid tail and hind limb weakness; 3, partial hind limb paralysis; 4, total hind limb paralysis; 5, hind and fore leg paralysis). OCH (400 μg/kg;) or vehicle (CMC) was administered orally at the indicated timepoints. For antibody treatment, anti-IL-4 (11B11, UltraLEAF, Biolegend) and anti-CD20 (SA271G2, UltraLEAF, Biolegend) were injected i.v. as noted at 400μg and 200 μg per mouse per dose respectively. Single cell suspensions were prepared from the spleen, spine and gut as described previously (*75*). For stimulation experiments, culture media used was DMEM supplemented with 10% FCS, 2 mM L-glutamine, 100 U/ml of penicillin-streptomycin, and 50 µM 2-Mercaptoethanol (all Invitrogen/Thermo).

### Soluble factor measurement

Cytokine concentrations in supernatants 96 hours after stimulation were measured by ELISA according to manufacturer’s instructions; OptEIA Set Immunoassay for IL-4, GM-CSF, and IFN-γ (BD Biosciences), and Duoset Immunoassay for IL-17 (R&D Systems, Minneapolis, MN, USA). Serum and supernatant antibody levels were measured using Mouse total IgG Ready-set-Go! ELISA kits (Thermo) according to manufacturer’s instructions.

### Statistical analyses

Data were analysed using a Prism software (GraphPad, Boston, MA, USA). Normality was assessed using a Shapiro-Wilk test. For direct before/after comparisons, Paired Student’s *t*-test or Wilcoxon matched-pairs signed rank test were used for normal or non-parametric data respectively. For comparison between two groups, an Unpaired Student’s *t*-test or Mann-Whitney *U* test were then used as appropriate with Welch’s correction when an F-test indicated unequal variances. For multiple group comparisons, a one-way or two-way ANOVA was performed as specified, followed by Tukey’s or Dunn’s multiple comparisons test. For contingency testing, a Fisher’s exact test was used and odds ratios (OR) were calculated. A *p* value < 0.05 was considered as significant. ns, not significant; *, *p* < 0.05; **, *p* < 0.01; ***, *p* < 0.001; ****, *p* < 0.0001.

## Data Availability

All data produced in the present study are available upon reasonable request to the authors

## Acknowledgements

We thank the NIH Tetramer Core Facility (contract number 75N93020D00005) for providing CD1d-PBS-57 tetramers. This research was supported by Japan Agency for Medical Research and Development (AMED) grant number JP18ek0109342h0001 and JSPS KAKENHI grants JP24K10278 and JP25K10802.

## Author Contributions

Conceptualization and Study design: TY, TO, RS, YA WS, SM, HN, NS

Investigation: BJER, AK, TO, YS, YN, T-Yo, YL, and WS

Data curation and Analysis: YL, YK, NS, YS, YN, TN, T-Yo, TI, HN, WS, TO, BJER, NM, AK, MT

Writing - Original Draft: BJER, AK, WS

Writing - Review & Editing, TY, WS, BJER, AK

Funding Acquisition: TY

Resources: MT, YO, TI, NM, and HN

Supervision: TY, WS

