## Supplementary Figures and Tables for "Sustained freedom from disease activity in secondary progressive multiple sclerosis by targeting invariant NKT cells: a phase 2 trial of OCH"

**Supplementary Materials**
Supplementary Figures 1-6
Supplementary Tables 1-9

**Supplementary Figure 1**
Adverse events (AEs) were collected throughout the study.
**(A)** Total AEs in each treatment group, stratified by severity grade: 1+ (all events, Grade 1-5), 2+ (Grade 2-5), 3+ (Grade 3-5).
**(B-D)** Distribution of AEs by System Organ Class (SOC) for Grade 1+ (B), Grade 2+ (C), and Grade 3+ (D).
SOC categories included: Infections and infestations; Blood and lymphatic system disorders; Psychiatric disorders; Metabolism and nutrition disorders; Nervous system disorders; Eye disorders; Ear and labyrinth disorders; Cardiac disorders; Vascular disorders; Respiratory, thoracic and mediastinal disorders; Gastrointestinal disorders; Skin and subcutaneous tissue disorders; Musculoskeletal and connective tissue disorders; Reproductive system and breast disorders; General disorders and administration site conditions; Investigations; Injury, poisoning and procedural complications.

**Supplementary Figure 2**
Patients were evaluated for number and volume of lesions at baseline and 24 weeks.
**(A)** Baseline lesion counts for patients stratified into four subgroups according to treatment assignment (placebo or OCH) and the presence or absence of new lesions during the study period.
**(B)** Changes in lesion volume from baseline to the end of the study shown by group (left) and longitudinally for individual patients (right).
**(C)** Reverse KM curves for time to first new lesion in patients treated with placebo or OCH, analyzed separately for relapsing-remitting multiple sclerosis (RRMS; left; placebo, n = 8; OCH, *n* = 8) and secondary progressive multiple sclerosis (SPMS; right; placebo, n = 6; OCH, n = 6).

**Supplementary Figure 3.**
(A) Change in EDSS (ΔEDSS) from baseline to end of study in OCH- versus placebo-treated patients, shown for all patients (left), RRMS (centre) and SPMS (right).
(B) Individual EDSS score trajectories over the study period for all patients (left), RRMS (centre) and SPMS (right).
(C) Change in Functional System scores (ΔFS) from baseline to end of study for OCH versus Placebo treatments, shown for all patients (left), RRMS (centre) and SPMS (right).
(D) Individual Functional System score trajectories over the study period for all patients (left),

RRMS (centre) and SPMS (right).
(E) Total brain volume (left), grey matter volume (center) and white matter volume (right) from baseline to end of study for OCH versus placebo in RRMS and SPMS patients.

**Supplementary Figure 4**
Blood was drawn on the day of induction prior to first drug administration and at 24 weeks at the end of the study. Peripheral blood mononuclear cells (PBMC) were isolated and restimulated *in vitro* for 4 hours before evaluation of marker expression and cytokine production by flow cytometry. Th cells were defined as CD3+CD56-CD4+CD8-.
(A) Comparison of IFN-γ producing Th cells between placebo and OCH-treated patients at baseline and 24 weeks for all cases (left) or divided by diagnosis, RRMS (center) and SPMS (right).
(B) Comparison of IL-17 producing Th cells between placebo and OCH-treated patients at baseline and 24 weeks for all cases (left) or divided by diagnosis, RRMS (center) and SPMS (right).
(C) GM-CSF+ Th cells over the study are shown for only cases that maintained NEDA status during the whole study.
Discontinued cases were excluded, placebo, n=12; OCH, n=13). For NEDA analysis, placebo, n=4; OCH, n=10)

**Supplementary Figure 5**
(A) EAE was induced in C57BL/6 mice by $MOG_{35-55}$ immunization with groups of mice receiving oral OCH or vehicle (CMC). At peak acute EAE, spleen or spinal-infiltrating Th cells (CD45+CD11b-TcRβ+CD4+CD8-) were isolated, restimulated with PMA/ionomycin, and assessed by intracellular flow cytometry for IFN-γ-producing Th cells (left) or IL-4-producing Th cells (right).
n = 4-5 per group, data are representative of 2-4 independent experiments.
(B) Acute EAE was induced in C57BL/6 mice by $MOG_{35-55}$ immunization and mice received weekly oral treatments either OCH dissolved in CMC and or CMC alone (vehicle). Immune cells were isolated from CNS at peak disease and restimulated with 2 μg/ml $MOG_{35-55}$ peptide. After 96 hour supernatants were harvested and assessed for the concentration of IFN-γ, IL-17 and GM-CSF by ELISA (B).
Data are from 2 independent experiments, with triplicates from pooled cell populations.
(C) Lymphocytes were isolated from spleen of mice at peak EAE treated either with OCH or vehicle by weekly oral doses or from or unmanipulated wildtype littermates (WT). CD19+TcRb- B cell subsets were assessed by flow cytometry as follows:
Follicular B cells (IgD+; left), MZ B cells (Marginal zone IgD-IgM+; center), and Class Switched B cells (IgM-IgD-; right)

(D) Plasmablasts (CD138+; left), GC B cells (Germinal center, GL7+; center), and CD69+ activated B cells (right)
n= 4-5 per group Data are representative of 3 independent experiments.
(E) Serum was harvested from mice at peak EAE treated either with OCH or vehicle by weekly oral doses or from or unmanipulated wildtype littermates (WT). IgG was measured by ELISA.
n= 5 per group Data are representative of 3 similar experiments.

**Supplementary Figure 6**
Serum analysis of 4 RRMS patients treated weekly with 0.3 mg oral OCH from the Step 2 section of our phase I trial (*19*). Antibody concentration in serum was measured for different isotypes by luminex. Plots show antibody class levels in serum for (left to right) (A) IgG1, IgG2, and IgG3 and (B) IgG4, IgA, IgM in 4 individual RRMS patients at baseline prior to OCH induction (0) and 4, 8, 12 weeks where samples were available. Time effect was assessed by RM-ANOVA.

**Supplementary Tables**

**Supplementary Table 1.**
Summary of cohort selection criteria, including inclusion and exclusion criteria and prohibited concomitant medications with minimum washout periods. Full details have been reported previously (*70*) and are registered under ClinicalTrials.gov (NCT04211740).

**Supplementary Table 2.**
Patient demography for the study cohort and baseline disease characteristics.

**Supplementary Table 3.**
List of patients withdrawn from the study. Treatment assignment (Placebo or OCH), diagnosis (RRMS or SPMS), duration of study participation prior to withdrawal (days of 180; percentage completed), and reason for discontinuation are shown. Reasons for discontinuation included recurrence of serious clinical disease (defined as two relapses during the trial or the requirement for three steroid pulses for a single relapse) or investigator decision (decline in lymphocyte count below enrolment criteria observed as baseline).

**Supplementary Table 4.**
List of serious adverse events observed during the trial period.

**Supplementary Table 5.**

Adverse events categorized by System Organ Class (SOC) and further subdivided by Primary Term (PT), with severity graded 1-5. Numbers indicate case-events, with the percentage of patients affected shown in parentheses. Table 5.1 corresponds to the placebo-treated group, and Table 5.2 corresponds to the OCH-treated group.

**Supplementary Table 6.**
Lesions were assessed at baseline, 3 months, and 6 months using gadolinium (Gd) contrast MRI by expert radiologists. The number of patients with new lesions at the end of the trial is summarized by treatment group for all enrolled participants. †Fisher's Exact Test.

**Supplementary Table 7.**
Clinical exacerbations were recorded by attending physicians during the study. Observer years were calculated based on the duration each participant was under treatment or observation. The annualized relapse rate (ARR) was determined by dividing the numbers of relapse by the observer years. The number of relapses in the two years prior to the study was obtained from medical records and categorized as those occurring 1-2 years before the study and those occurring in the year immediately preceding the study. Data are summarized by treatment group for all enrolled participants (placebo, n = 15; OCH, n = 15).

**Supplementary Table 8**
Adverse events were recorded by attending physicians during the study based on 1 or more protocol-defined recurrences, or increase in functional impairment confirmed via EDSS or FS, or at least 1 MRI change (contrast-enhanced lesion on T1-3D CE imaging or new or enlarged preexisting lesion on T2-weighted imaging). Data are summarized by group for all enrolled cases (placebo, n = 15; OCH, n = 15). †Fisher's Exact Test

**Supplementary Table 9**
List of antibodies used in the study.

Supplementary Table 1

### Inclusion criteria: Main

- Patients aged 20-65 years who provided written consent for trial participation
- RRMS or SPMS according to revised McDonald criteria as recorded in prior medical record
- Relapsing: 2 clinical exacerbations within previous 24 months or 1 exacerbation within previous 12 months

### Inclusion criteria: Screening

- At least one lesion suspected to be MS on screening MRI
- EDSS = 7
- No clinical or test findings suggesting acute recurrence based on evaluation by a neurologist
- Peripheral blood lymphocyte count <600/$mm^3$

### Exclusion criteria: Screening

- NMOSD diagnosis including anti-aquaporin-4 antibody-positive
- Currently pregnant or nursing
- Contraindication for MRI or sensitivity to gadolinium contrast
- Immunocompromised patients or patients with suspected infectious disease, or patients with Inflammatory bowel disease, Liver dysfunction or history of liver disease, or malignant tumors (previous 5 years)
- Varicella-zoster virus IgG antibody–negative
- Positive for syphilis serum reaction, β-D glucan, T-spot test, human immunodeficiency virus infection, hepatitis B infection
- History of stem cell transplantation, organ transplantation, and treatment for rejection
- Physical, mental, or social condition affecting the ability to provide consent to or complete the trial
- Blood donation (200 mL within 2 months, 400 mL within 3 months) prior to enrolment

### Exclusion criteria: previous/concomitant therapies:

| Drug | Prohibited period |
|---|---|
| Interferon-β preparation | 1 month before trial enrolment |
| Fingolimod hydrochloride | 6 months before trial enrolment |
| Natalizumab | 3 months before trial enrolment |
| Glatiramer Acetate | 1 month before trial enrolment |
| Dimethyl Fumarate | 3 months before trial enrolment |
| Immunosuppressive drugs (except azathioprine) with immunostimulatory/myelosuppressive effects | 3 months before trial enrolment |
| Pulse therapy using corticosteroids | 1 month before trial enrolment (except when MS recurs); |
| Plasmapheresis<br>Immunoadsorption therapy<br>Lymphocyte depletion therapy | Prohibited from the time of consent |
| Immunoglobulin preparation | 2 months before trial enrolment |
| Vaccines | 1 month before trial enrolment |
| Other investigational drugs/Clinical trials | 4 months before trial enrolment |

### Additional exclusion criteria:

- Abnormal cardiac indictors: ECG, a history of other risk factors for torsades de pointes
- History of severe drug or food allergy, history of seizures/epilepsy, or Asthma (previous 10 years)
- Patients with drug or alcohol dependence in the past or present)
- Other pathological symptoms, illnesses, or history that may affect this trial

Supplementary Table 2

| | All | Placebo | OCH |
|---|---|---|---|
| Number of patients | 30 | 18 | 12 |
| Male (%) | 10 (33.3) | 5 (33.3) | 5 (33.3) |
| Female (%) | 20 (66.6) | 10 (66.6) | 10 (66.6) |
| RRMS | 18 | 9 | 9 |
| SPMS | 12 | 6 | 6 |
| Mean Age; years (SD) | 45.0 (9.5) | 46.5 (9.0) | 43.4 (10.0) |
| Mean Age at onset; years (SD) | 33.2 (9.7) | 34.2 (9.1) | 32.2 10.6) |
| Mean Disease Duration; Years (SD) | 13.5 (12.3) | 14.07 (11.3) | 12.9 (13.5) |
| Median Disease Duration; Years (inter-quartile) | 11 (19) | 15 (18) | 11 (17) |
| Mean EDSS (SD) | 4.2 (2.1) | 3.9 (2.2) | 4.4 (2.0) |
| Average relapses, prior 2 years | 2.47 (1.17) | 2.33 (1.35) | 2.60 (0.99) |

| Patient | Drug | Diagnosis | Study length | % Completed | Discontinuation Reason |
|---|---|---|---|---|---|
| OCH-05 | Placebo | RRMS | 127/180 | 71% | 2 relapses |
| OCH-11 | Placebo | RRMS | 63/180 | 35% | Low baseline lymphocytes |
| OCH-10 | OCH | RRMS | 71/180 | 39% | Low baseline lymphocytes |
| OCH-29 | OCH | RRMS | 96/180 | 53% | 2 relpases |
| OCH-15 | Placebo | SPMS | 34/180 | 19% | 3 steroid pulses |

Supplementary Table 3: Summary of patients exiting trial

| Patient | Drug | Onset day | Duration (days) | Adverse Event |
|---|---|---|---|---|
| OCH-07 | Placebo | 136/183 | 10 | Gastroenteritis (Grade 3) |

Supplementary Table 4  Serious Adverse Events

Supplementary Table 5.1 Adverse Event Severity by SOC and PT, SS (Placebo treatment group)

| System Organ Class (SOC)/Primary Term (PT) | Placebo group (n=15) | | | | |
|---|---|---|---|---|---|
| | Grade 1 | Grade 2 | Grade 3 | Grade 4 | Grade 5 |
| Number of cases | 0 (0.0) | 9 (60.0) | 1 (6.7) | 0 (0.0) | 0 (0.0) |
| Infectious Diseases and Parasitic Diseases | 0 | 2 (13.3) | 1 (6.7) | 0 | 0 |
| Gastroenteritis | 0 | 0 | 1 (6.7) | 0 | 0 |
| Pharyngitis | 0 | 1 (6.7) | 0 | 0 | 0 |
| Pneumonia | 0 | 1 (6.7) | 0 | 0 | 0 |
| Metabolic and Nutritional Disorders | 0 | 1 (6.7) | 0 | 0 | 0 |
| Hypoglycemia | 0 | 1 (6.7) | 0 | 0 | 0 |
| Nervous system disorders | 1 (6.7) | 2 (13.3) | 0 | 0 | 0 |
| Floating dizziness | 1 (6.7) | 1 (6.7) | 0 | 0 | 0 |
| Headache | 1 (6.7) | 1 (6.7) | 0 | 0 | 0 |
| Somnolence | 1 (6.7) | 0 | 0 | 0 | 0 |
| Eye disorders | 0 | 3 (20.0) | 0 | 0 | 0 |
| Conjunctival deposits | 0 | 1 (6.7) | 0 | 0 | 0 |
| Eye pain | 0 | 1 (6.7) | 0 | 0 | 0 |
| Foreign body sensation in the eye | 0 | 1 (6.7) | 0 | 0 | 0 |
| Ear and labyrinth disorders | 0 | 1 (6.7) | 0 | 0 | 0 |
| Rotatory vertigo | 0 | 1 (6.7) | 0 | 0 | 0 |
| Heart disorders | 1 (6.7) | 0 | 0 | 0 | 0 |
| Bradycardia | 1 (6.7) | 0 | 0 | 0 | 0 |
| Vascular disorders | 1 (6.7) | 0 | 0 | 0 | 0 |
| Vascular disorders | 1 (6.7) | 0 | 0 | 0 | 0 |
| Gastrointestinal disorders | 0 | 1 (6.7) | 0 | 0 | 0 |
| Diarrhea | 0 | 1 (6.7) | 0 | 0 | 0 |
| Skin and subcutaneous tissue disorders | 1 (6.7) | 2 (13.3) | 0 | 0 | 0 |
| Hives | 1 (6.7) | 1 (6.7) | 0 | 0 | 0 |
| Alopecia | 1 (6.7) | 0 | 0 | 0 | 0 |
| Eczema | 0 | 1 (6.7) | 0 | 0 | 0 |
| Rash | 0 | 1 (6.7) | 0 | 0 | 0 |
| Musculoskeletal and connective tissue disorders | 0 | 3 (20.0) | 0 | 0 | 0 |
| Joint pain | 0 | 1 (6.7) | 0 | 0 | 0 |
| Neck pain | 0 | 1 (6.7) | 0 | 0 | 0 |
| Herniated disc | 0 | 1 (6.7) | 0 | 0 | 0 |
| Reproductive System and Breast Disorders | 0 | 1 (6.7) | 0 | 0 | 0 |
| Dysmenorrhea | 0 | 1 (6.7) | 0 | 0 | 0 |

Supplementary Table 5.1 (Continued)

| System Organ Class (SOC)/Primary Tumor (PT) | Placebo group (n=15) | | | | |
|---|---|---|---|---|---|
| | Grade 1 | Grade 2 | Grade 3 | Grade 4 | Grade 5 |
| General/Systemic disorders and administration site conditions | 1 (6.7) | 4 (26.7) | 0 | 0 | 0 |
| Fever | 0 | 2 (13.3) | 0 | 0 | 0 |
| Chest pain | 1 (6.7) | 0 | 0 | 0 | 0 |
| Condition Aggravated | 0 | 1 (6.7) | 0 | 0 | 0 |
| Edema | 0 | 1 (6.7) | 0 | 0 | 0 |
| Clinical Laboratory | 1 (6.7) | 1 (6.7) | 0 | 0 | 0 |
| Increased blood creatine phosphokinase | 0 | 1 (6.7) | 0 | 0 | 0 |
| Leukocytosis | 1 (6.7) | 0 | 0 | 0 | 0 |
| Proteinuria | 1 (6.7) | 0 | 0 | 0 | 0 |
| Injuries, Poisonings, and Treatment Complications | 0 | 2 (13.3) | 0 | 0 | 0 |
| Frostbite | 0 | 1 (6.7) | 0 | 0 | 0 |
| Fall | 0 | 1 (6.7) | 0 | 0 | 0 |
| Skin laceration | 0 | 1 (6.7) | 0 | 0 | 0 |

MedDRA/J Version 24.1

Supplementary Table 5.2 Adverse Event Severity by SOC and PT, SS (OCH treatment group)

| System Organ Class (SOC)/Primary Term (PT) | OCH-NCNP1 Group (n=15) | | | | |
|---|---|---|---|---|---|
| | Grade 1 | Grade 2 | Grade 3 | Grade 4 | Grade 5 |
| Number of cases | 6 (40.0) | 5 (33.3) | 2 (13.3) | 0 (0.0) | 0 (0.0) |
| Infectious Diseases and Parasitic Diseases | 0 | 3 (20.0) | 0 | 0 | 0 |
| Cystitis | 0 | 1 (6.7) | 0 | 0 | 0 |
| Gingivitis | 0 | 1 (6.7) | 0 | 0 | 0 |
| Nasopharyngitis | 0 | 1 (6.7) | 0 | 0 | 0 |
| Blood and lymphatic system disorders | 0 | 0 | 1 (6.7) | 0 | 0 |
| Lymphopenia | 0 | 0 | 1 (6.7) | 0 | 0 |
| Mental disorders | 0 | 1 (6.7) | 0 | 0 | 0 |
| Insomnia | 0 | 1 (6.7) | 0 | 0 | 0 |
| Nervous system disorders | 2 (13.3) | 1 (6.7) | 0 | 0 | 0 |
| Headache | 1 (6.7) | 1 (6.7) | 0 | 0 | 0 |
| Floating dizziness | 1 (6.7) | 0 | 0 | 0 | 0 |
| Eye disorders | 0 | 1 (6.7) | 0 | 0 | 0 |
| Eyelid swelling | 0 | 1 (6.7) | 0 | 0 | 0 |
| Heart disorders | 3 (20.0) | 0 | 0 | 0 | 0 |
| Palpitations | 1 (6.7) | 0 | 0 | 0 | 0 |
| Sinus bradycardia | 1 (6.7) | 0 | 0 | 0 | 0 |
| Supraventricular Premature Contraction | 1 (6.7) | 0 | 0 | 0 | 0 |
| Respiratory, thoracic, and mediastinal disorders | 1 (6.7) | 1 (6.7) | 0 | 0 | 0 |
| Upper respiratory tract inflammation | 1 (6.7) | 0 | 0 | 0 | 0 |
| Cough-variant asthma | 0 | 1 (6.7) | 0 | 0 | 0 |
| Gastrointestinal disorders | 3 (20.0) | 2 (13.3) | 0 | 0 | 0 |
| Abdominal discomfort | 1 (6.7) | 0 | 0 | 0 | 0 |
| Upper abdominal pain | 1 (6.7) | 0 | 0 | 0 | 0 |
| Angular Cheilitis | 1 (6.7) | 0 | 0 | 0 | 0 |
| Diarrhea | 1 (6.7) | 0 | 0 | 0 | 0 |
| Gastritis | 0 | 1 (6.7) | 0 | 0 | 0 |
| Vomiting | 0 | 1 (6.7) | 0 | 0 | 0 |
| Skin and subcutaneous tissue disorders | 1 (6.7) | 2 (13.3) | 1 (6.7) | 0 | 0 |
| Hypohidrosis | 0 | 0 | 1 (6.7) | 0 | 0 |
| Rash | 0 | 1 (6.7) | 0 | 0 | 0 |
| Skin discoloration | 1 (6.7) | 0 | 0 | 0 | 0 |
| Hives | 0 | 1 (6.7) | 0 | 0 | 0 |
| Musculoskeletal and connective tissue disorders | 2 (13.3) | 1 (6.7) | 0 | 0 | 0 |
| Back pain | 0 | 1 (6.7) | 0 | 0 | 0 |
| Joint swelling | 1 (6.7) | 0 | 0 | 0 | 0 |
| Limb pain | 1 (6.7) | 0 | 0 | 0 | 0 |

Supplementary Table 5.2 (Continued)

| System Organ Class (SOC)/Primary Term (PT) | OCH-NCNP1 Group (n=15) Grade 1 | Grade 2 | Grade 3 | Grade 4 | Grade 5 |
|---|---|---|---|---|---|
| General/Systemic disorders and administration site conditions | 5 (33.3) | 0 | 0 | 0 | 0 |
| Impotence | 1 (6.7) | 0 | 0 | 0 | 0 |
| Chest pain | 1 (6.7) | 0 | 0 | 0 | 0 |
| Chills | 1 (6.7) | 0 | 0 | 0 | 0 |
| Facial edema | 1 (6.7) | 0 | 0 | 0 | 0 |
| Fever | 1 (6.7) | 0 | 0 | 0 | 0 |
| Clinical Laboratory | 1 (6.7) | 0 | 0 | 0 | 0 |
| ECG Abnormal T Wave | 1 (6.7) | 0 | 0 | 0 | 0 |
| Injuries, Poisonings, and Treatment Complications | 3 (20.0) | 1 (6.7) | 0 | 0 | 0 |
| Fall | 1 (6.7) | 0 | 0 | 0 | 0 |
| Muscle injury | 0 | 1 (6.7) | 0 | 0 | 0 |
| Contusion | 1 (6.7) | 0 | 0 | 0 | 0 |
| Post-treatment pruritus | 1 (6.7) | 0 | 0 | 0 | 0 |

MedDRA/J Version 24.1

| Dosing Period | | Placebo (%) | OCH (%) | Total (%) | Difference between groups | p value † | 95% CI |
|---|---|---|---|---|---|---|---|
| 0-12 weeks | No new lesions | 12 (92.3) | 13 (92.9) | 25 (92.6) | -0.5 | p=1.000 | -29.5-27.2 |
| | New lesions | 1 (7.7) | 1 (7.1) | 2 (7.4) | | | |
| | Total | 13 | 14 | 27 | | | |
| 12-24 weeks | No new lesions | 10 (83.3) | 13 (100.0) | 23 (92.0) | -16.7 | p=0.220 | -48.4-9.8 |
| | New lesions | 2 (16.7) | 0 (0.0) | 2 (8.0) | | | |
| | Total | 13 | 12 | 25 | | | |
| 0-24 weeks (Total period) | No new lesions | 14 (93.3) | 12 (80.0) | 26 (86.7) | -13.3 | p=0.598 | -43.3-14.9 |
| | New lesions | 1 (6.7) | 3 (20.0) | 4 (13.3) | | | |
| | Total | 15 | 15 | 30 | | | |

Supplementary Table 6: Appearance of New Lesions (MRI)

| | | Placebo n=15 | OCH n=15 | Total n=30 |
|---|---|---|---|---|
| Study period | Number of Relapses | 8 | 4 | 12 |
| | Observer Year | 6.825 | 7.075 | 13.900 |
| | Annual relapse rate (ARR /person-years) | 1.172 | 0.565 | 0.863 |
| Prior to study period | Number of Relapses (1-2 years previous) | 15 | 22 | 37 |
| | Number of Relapses (previous 1 year) | 18 | 15 | 32 |
| | ARR | 1.167 | 1.300 | 1.233 |

Supplementary Table 7 Annualised relapse rate

| Dosing Period | | Placebo (%) | OCH (%) | Total (%) | Difference between groups | p value † | 95% CI |
|---|---|---|---|---|---|---|---|
| 0-24 weeks (Total period) | No events | 5 (33.3) | 10 (66.7) | 15 (50) | -33.3 | p=0.143 | -65.4-5.6 |
| | Adverse Event | 10 (66.7) | 5 (33.3) | 15 (50) | | | |
| | Total | 15 | 15 | 30 | | | |

Supplementary Table 8: NEDA – no evidence of disease activity

| Target | Species | Fluorochrome | Manufacturer | Catalogue Number |
|---|---|---|---|---|
| CD161 | Human | PerCP-Cy5.5 | BioLegend | 339908 |
| CD3 | Human | V500 | BD Horizon | 561416 |
| CD4 | Human | APC-H7 | BD | 560158 |
| CD56 | Human | BV421 | BioLegend | 318328 |
| CD8 | Human | ECD | Beckman Coulter | 737659 |
| CD8β | Human | ECD | Beckman Coulter | 6607123 |
| αGalCer-CD1d tetramer | Human | APC | NIH Tetramer Core Facility | PBS-57-APC |
| γδTCR | Human | FITC | BioLegend | 331208 |
| GM-CSF | Human | PE | BioLegend | 502306 |
| IFN-γ | Human | PerCP-Cy5.5 | BioLegend | 502526 |
| IL-10 | Human | PE | BD Pharmingen | 559330 |
| IL-17A | Human | Alexa488 | BioLegend | 512308 |
| IL-4 | Human | APC | BioLegend | 500714 |
| IL-5 | Human | PE | BioLegend | 500904 |
| m IgG1 k | Human | Alexa488 | BioLegend | 400134 |
| m IgG1 k | Human | APC | BioLegend | 400120 |
| m IgG1 k | Human | PerCP-Cy5.5 | BioLegend | 400150 |
| rat IgG2a k | Human | PE | BD Pharmingen | 554689 |
| TCR Vα7.2 | Human | PE | BioLegend | 351706 |
| CD45 | Mouse | BV510 | BioLegend | 103137 |
| TcRb | Mouse | PE-Cy7 | BioLegend | 109222 |
| CD4 | Mouse | PerCP-Cy5.5 | BioLegend | 100434 |
| CD8 | Mouse | FITC | BioLegend | 100706 |
| CD11b | Mouse | APC-Fire750 | BioLegend | 101262 |
| IFN-g | Mouse | BV421 | BioLegend | 505829 |
| IL-17 | Mouse | APC | BioLegend | 506916 |
| GM-CSF | Mouse | PE | BioLegend | 505406 |

Supplementary Table 9: Antibodies used in this study

Supplementary Figure 1

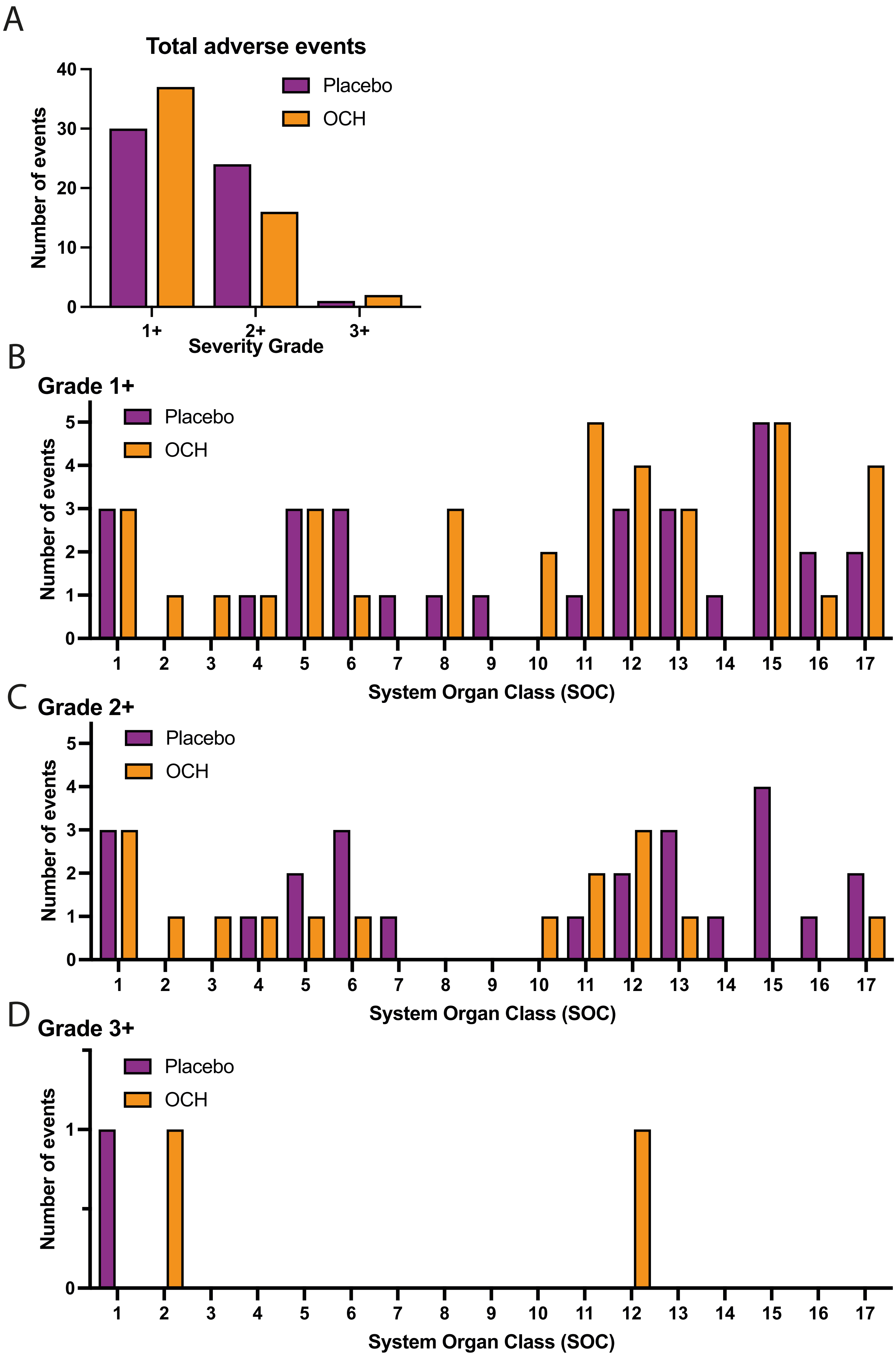

Supplementary Figure 2

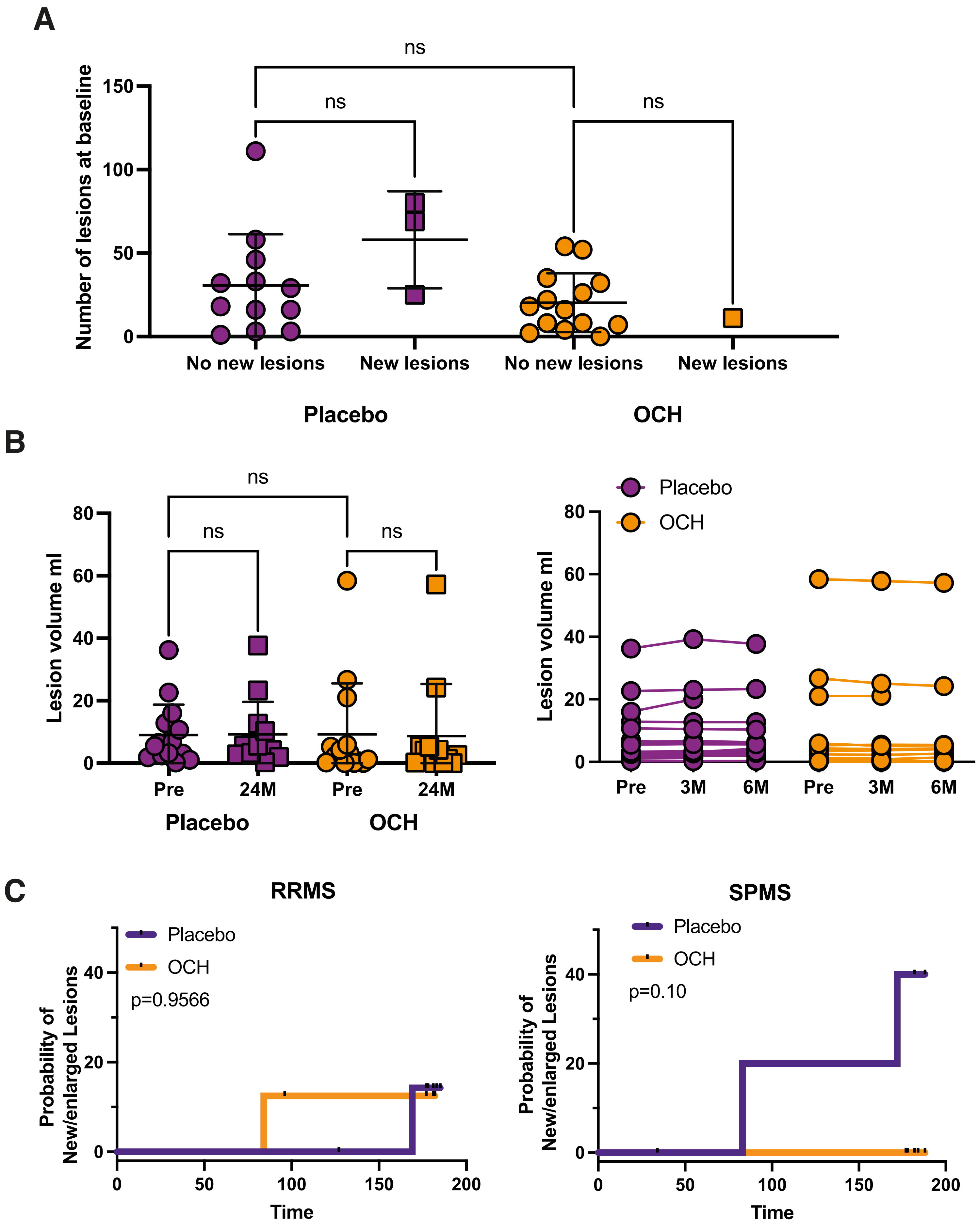

Supplementary Figure 3

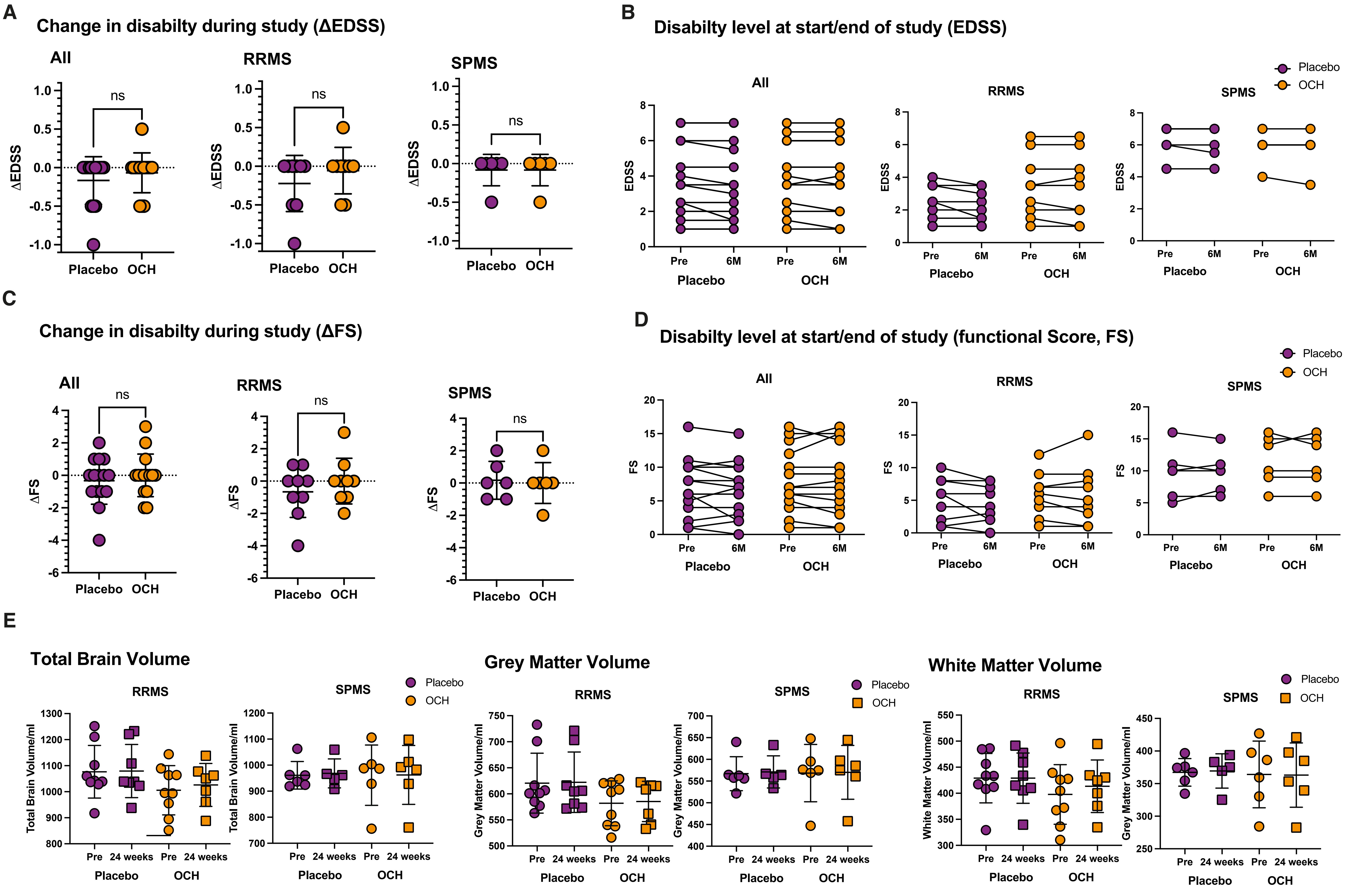

Supplementary Figure 4

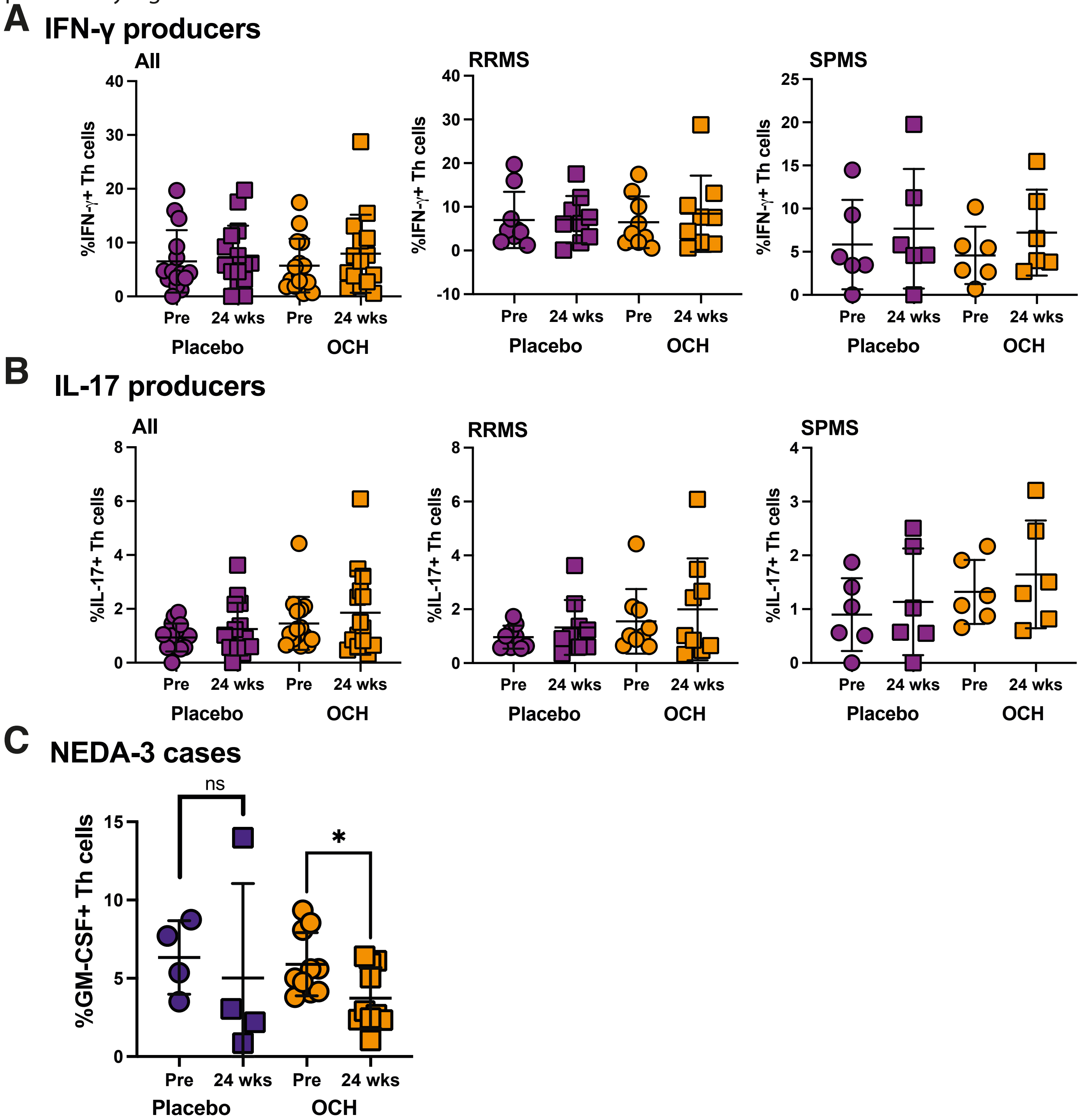

Supplementary Figure 5

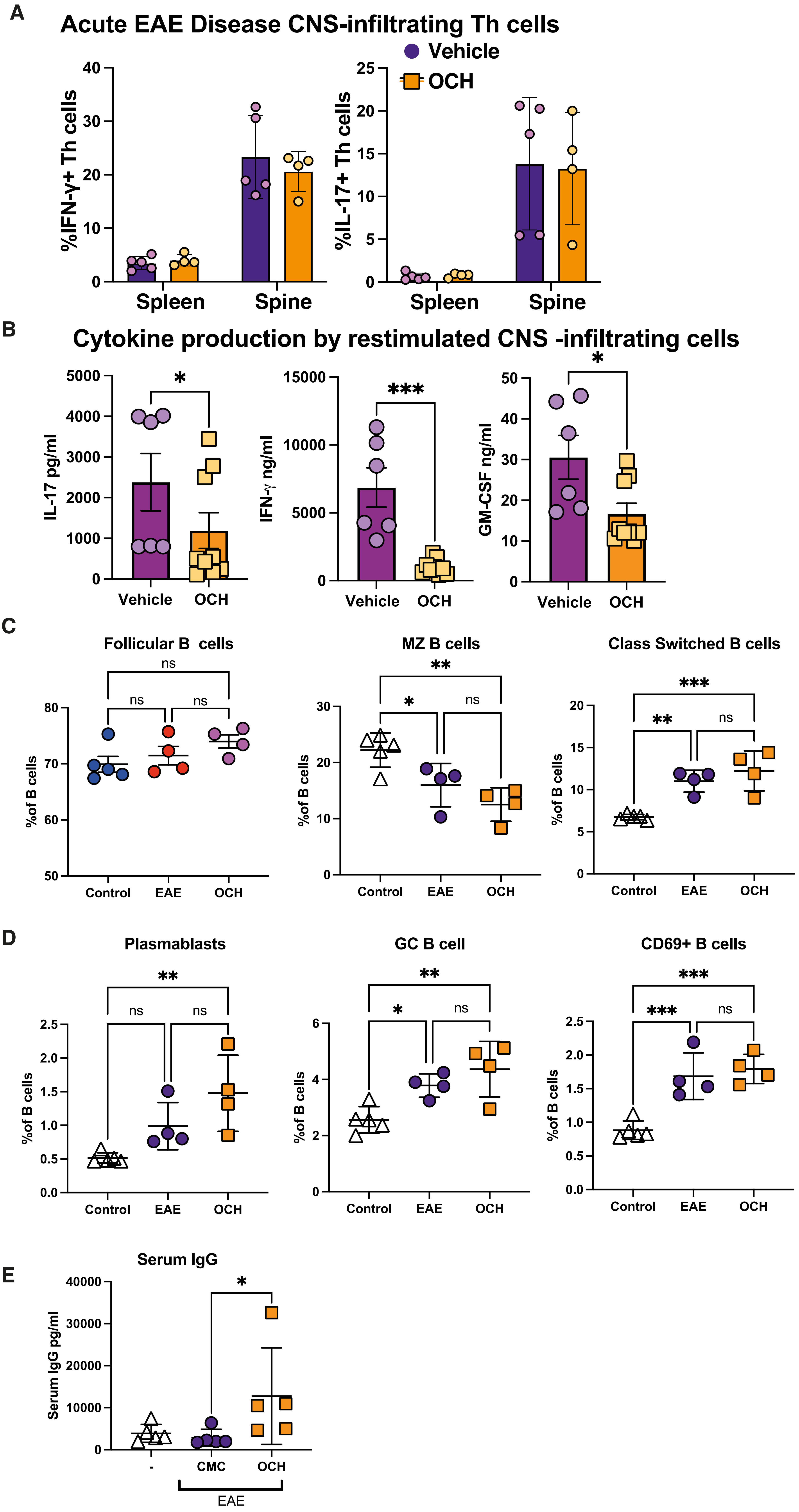

Supplementary Figure 6

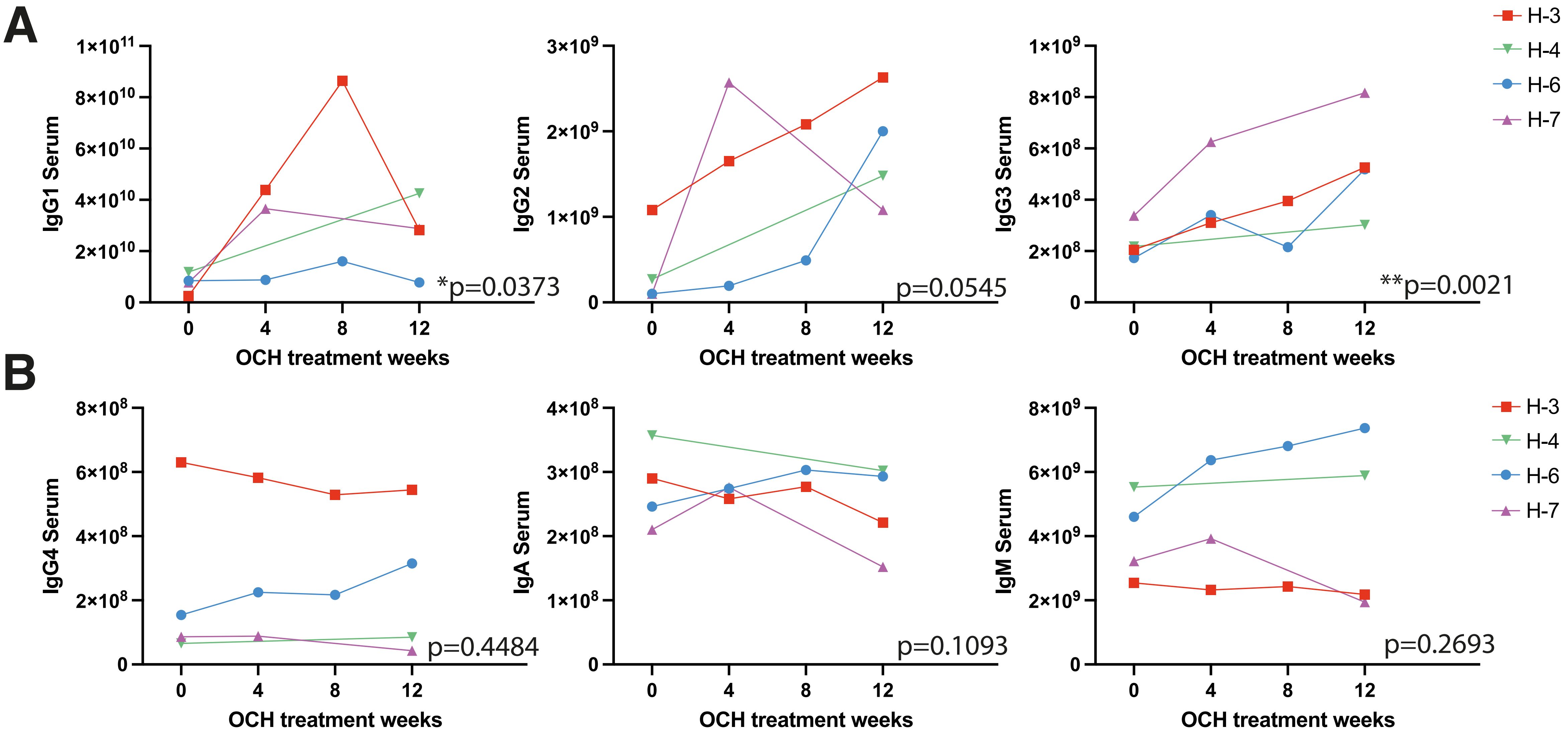
